# Effective post-exposure prophylaxis of Covid-19 associated with hydroxychloroquine: Prospective dataset re-analysis incorporating novel, missing data

**DOI:** 10.1101/2020.11.29.20235218

**Authors:** David M. Wiseman, Pierre Kory, Samir A Saidi, Dan Mazzucco

**Affiliations:** Synechion, Inc., Dallas, TX, USA; Aurora St. Luke’s Medical Center, Milwaukee, WI, USA. and Front Line Critical Care Alliance.; Central Clinic School, University of Sydney, Australia; Third Eye Associates, Cherry Hill, NJ, USA.

**Keywords:** COVID-19, Post-exposure prophylaxis (PEP), Hydroxychloroquine (HCQ), Re-analysis, SARS-Cov2

## Abstract

**BACKGROUND:** A key trial (NCT04308668) of post-exposure prophylaxis found hydroxychloroquine-associated (HCQ) reductions of Covid-19 by 17% overall and 31% to 49% in subgroups. To understand these trends, we re-analyzed the dataset.

**METHODS:** Our protocol conformed to the Standard Protocol Items: Recommendations for Interventional Trials (SPIRIT). We compared the incidence of Covid-19 after HCQ or placebo, stratifying by intervention lag, age, and gender.

**RESULTS:** Newly requested data missing from the dataset revealed that 52% and 19% of subjects received medication 1-2 days after intended and assumed overnight delivery or four-day intervention lag respectively. After re-analysis, we found reduced HCQ-associated incidence of Covid-19 with Early (up to 3 days post-exposure) (RR 0.58, 95%CI 0.35 - 0.97; *p*=0.044; NNT 14.5) but not Late (RR 1.22, 95%CI 0.72 - 2.04) prophylaxis.

We found a significant HCQ-associated Covid-19 reduction in subjects 18 to 45 years old with Early (RR 0.54, 95%CI 0.29-0.97; *p*=0.0448, NNT 11.5) but not Late (RR 1.02, 95%CI 0.55-1.89) prophylaxis, attenuated in older subjects (RR 0.75, 95%CI 0-27-2.05) and by co-morbidities. Although, we did not detect effects of gender, folate, zinc, or ascorbate, confounding effects cannot be excluded.

**CONCLUSIONS:** Using novel data and prospective re-analysis, hydroxychloroquine, in an age-dependent manner, was associated with reduced Covid-19 compatible illness when supplied for post-exposure prophylaxis between 1 and 3 days after high- or moderate-risk exposure, at higher loading and maintenance doses than in similar studies. The original study conclusions are controverted, and our finding warrants prospective confirmation.

Protocol registered at Open Science Framework: osf.io/fqtnw (last revised September 27, 2020,

**Highlights:**

- Missing data integrated with dataset re-analysis reversed findings of original study
- Hydroxychloroquine associated reduction (42%) of Covid-19 compatible illness found
- Effect in Post-exposure Prophylaxis when received 1-3 days after exposure
- Risk Ratio 0.58 (95% CI 0.35-0.97, p=0.044, NNT14.5)
- Findings controvert the conclusions of original study

## 1 Introduction

There have been (June 30 2021) around 181 million cases of Covid-19 and 3.9 million deaths worldwide, [1] about 15-20% of them within the USA. With early interest in deploying hydroxychloroquine (HCQ), the US Food and Drug Administration (FDA) issued an Emergency Use Authorization (EUA) in March 2020. [2] Lacking randomized clinical trial (RCT) data, emerging observational reports (with exceptions [3]) disfavored HCQ. With safety concerns, FDA cautioned [4] using HCQ outside hospital or trial settings on April 24, 2020.

HCQ became highly controversial, with suggestions that *“to some extent the media and social forces — rather than medical evidence — are driving clinical decisions and the global Covid-19 research agenda*.*”* [5] Against this background, on June 15, 2020 FDA revoked [2] HCQ’s EUA, citing only one just-published RCT addressing hospitalized patients [6] and only one [7] addressing prophylaxis. This latter study examined post-exposure prophylaxis (the “PEP” study) in 821 asymptomatic adults with exposure to Covid-19. Subjects received HCQ (1.4g first day, then 600 mg daily for 4 more days) or placebo (folate -USA; lactose -Canada). The study concluded that *“…HCQ did not prevent illness* […] *when initiated within 4 days after* […] *exposure”* (RR 0.83, 95%CI 0.58-1.18, p=0.35).

We [8] and others have criticized the study’s interpretation. Since this was a pragmatic trial, with typically greater heterogeneity and smaller effect sizes than in an explanatory trial, [9] powering the study for a 50% effect size appears over-ambitious, especially given its early termination. [10] Type II errors abound, and conclusions as to HCQ’s ineffectiveness cannot be drawn as the study was not powered as an equivalence study. A reduction of 17% is arguably [11] clinically meaningful. Rather than targeting specific clinical goals, the authors suggested [12] that the study was powered to collect data quickly under pandemic conditions.

Non-statistically significant signals of HCQ-associated efficacy included a 31% reduction among household cohabitees. There were age-dependent reductions found in other analyses [13] to be statistically significant. The folate placebo and ex-protocol use of zinc and ascorbate may have been confounding (Supplement). With a reduction of 49% associated with early (“Day 1”) HCQ prophylaxis, we [8] and others [14] found a negative association between intervention lag and effect.

We conjectured that *post hoc* exploratory re-analysis of the PEP study would inform a time-and age-nuanced approach to Covid-19 using HCQ, testable prospectively. Our objectives were to define: (a) time-or (b) age-dependent effects associated with HCQ and, (c) the influence of gender, exposure type, zinc, ascorbate or folate on outcomes.

## 2 Methods

### 2.1 Dataset and Protocol Revisions

One protocol (NCT04308668) described separately reported PEP [7] or early post-exposure treatment (PET) [15] cohorts. The de-identified PEP dataset was released (covidpep.umn.edu/data) with revisions (see Supplement for details): September 9 (“9/9”), October 6 (“10/6”) and October 30 (“10/30”) 2020. Another, inaccurate, version was released on October 26 (“10/26”).

Using the Open Science Framework (OSF) protocol template (osf.io/jea94/), we conformed to the SPIRIT checklist (Standard Protocol Items: Recommendations for Trials [16]) integrating the WHO Trial Registration Data Set. [17] Our protocol was registered on August 13, 2020 with revisions (Supplement), most recently September 27, 2020 (osf.io/vz8a7/ [8]) prior to receiving data (10/6 revision) regarding the time to drug receipt.

Four main areas required clarification (Supplement): (i) exposure risk definition; (ii) study medication adherence; (iii) “intervention lag” (time from exposure to receipt of medication, resolved by the 10/6 revision); (iv) nomenclature for timing study events. The authors notified us that the nomenclature implied by the use of the term “days” in their paper was incorrect. Although “Days from exposure to enrollment” (implying elapsed time) was stated (Table 1 of [7]), “days” (1 day, 2 days etc.) denoted the “Day on which enrollment occurred.” This yields confusing inconsistencies. It is important therefore to distinguish between the “Days from” nomenclature (elapsed time from exposure to drug receipt) and the “Day on” nomenclature (the numbered day on which drug was received -Day 2, Day 3 etc., with Day 1 as the day of highest-risk exposure, see Table 1).To our knowledge this has neither been corrected in the original nor in a subsequent work. [18]

**Table 1.**
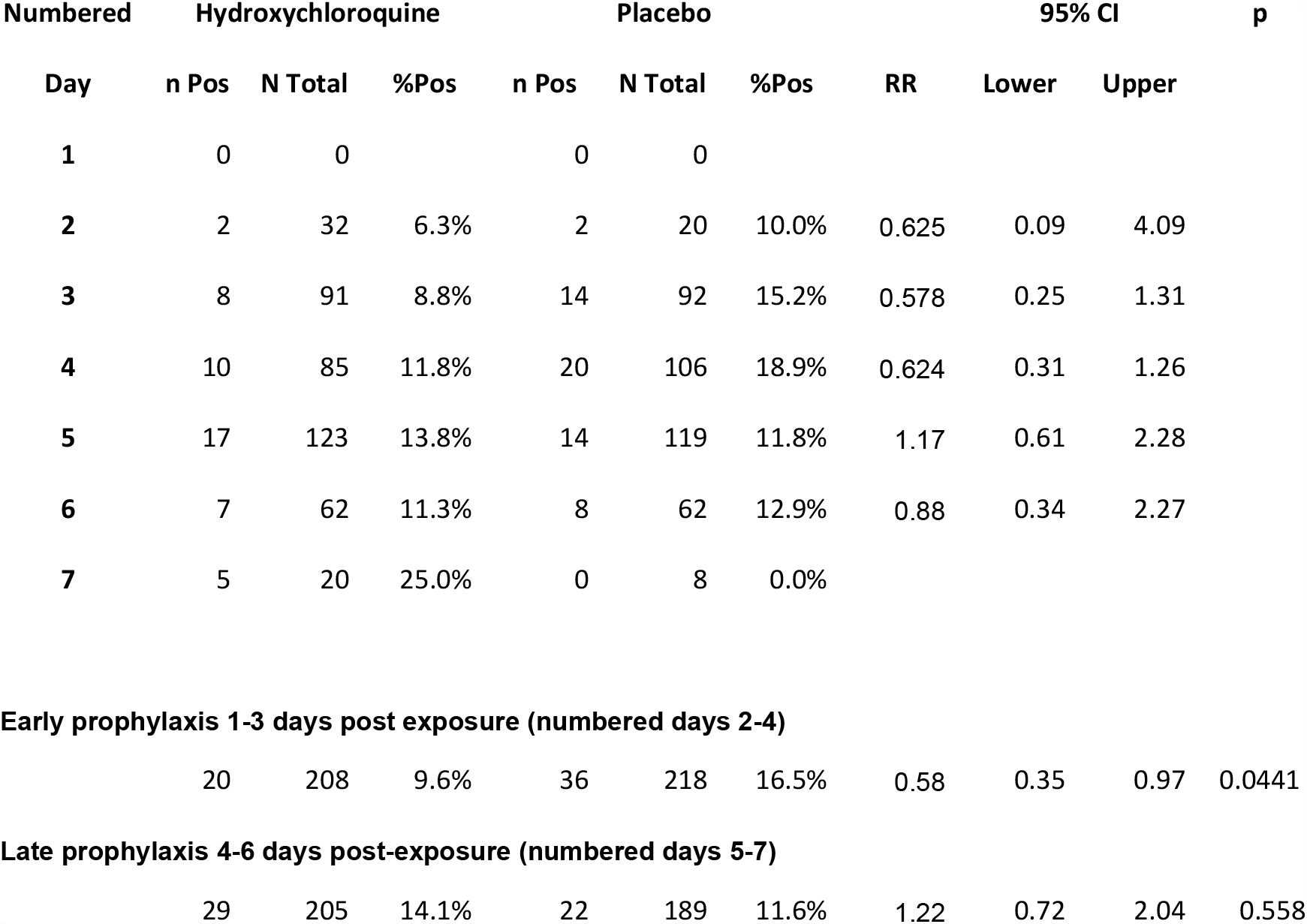
Stratification of effect associated with hydroxychloroquine based on time from exposure to drug receipt (ITT population) The number (and percent) of subjects with a Covid-19 positive outcome are shown for each group along with the total number of subjects for that group, stratified by time from exposure to drug receipt. Note there was one subject with missing data in the HCQ group. The upper part of the table uses the variable “exposure_days_to_drugstart” provided in the 10/6 dataset. As clarified by the original authors, “days” does not represent elapsed time (“Days from” nomenclature), but the numbered day on which study drug was received (“Day on” nomenclature), starting with “Day 1” as the day of reported high risk exposure. No patients received drug on the same day of exposure (i.e. “Day 1”). This terminology is used to allow comparison with Table 1 of the original publication which describes the time of enrollment. After the authors provided this clarification to us, it became evident that their original Table 1 mistakenly gives the impression that it is describing elapsed time from exposure (“Days from”) when in fact it is stratifying by the numbered day on (“Day on”) which enrollment occurred, with Day 1 being the day of high-risk exposure. To obtain elapsed time from exposure, 1 day must be subtracted from the numbered day. Accordingly, data for numbered days 2-4 and 5-7, respectively, have been aggregated in the lower part of the table into Early (1-3 days) and Late (4-6 days) cohorts, where “days” here describe elapsed time from exposure.

### 2.2 Analysis Plan

We re-stratified data by intervention lag followed by age, gender, exposure type, risk level, or use of zinc or ascorbate. An Intent-to-Treat (ITT) analysis was employed as in the original study. We analyzed data according to adherence to taking study medication, provision of outcome data, use of the folate placebo, and presence of co-morbidities (Supplement). We retained the original primary outcome variable: *“incidence of either laboratory-confirmed Covid-19 or illness compatible with Covid-19 within 14 days,”* comparing treatment arms using Fisher’s Exact test. We examined the severity of symptoms at 14 days according to a visual analogue scale originally described as a secondary outcome (Kruskal-Wallis test).

We mirrored the original use of two-tailed tests without adjustment for multiple comparisons. This is further justified by the exploratory nature of our analyses. p-values ≤ 0.05 were considered statistically significant. Larger values are presented to identify trends. Microsoft Excel was used for data processing and Vassar Stats (vassarstats.net/) for verification. The original authors provided calculations from which we verified our primary time stratification (Supplement). Extensive quality checks ensured the accuracy of the various datasets versions as well as our understanding of the data (Supplement). The original authors were invited to review our calculations, comment on and participate in this manuscript.

### 2.3 Ethics Committee Approval

No ethics committee approval was required as we used a de-identified, public dataset.

## 3 Results

### 3.1 Assumed Overnight Delivery vs. Actual Delivery Time

The original study had assumed that study drug sent by overnight shipping would be delivered overnight. We estimated [8] that within each of the reported [7] strata for “Time from exposure to enrollment” (1 to 4 days, incorrectly using the “Days from” rather than the “Day on” terminology), there could be overlapping variations in intervention lag, with for example subjects in the “1 day” or “4 days” strata receiving drug after the same interval. We requested from shipping-time derived data, missing from the original analysis and dataset, that would allow calculation of a more accurate intervention lag.

New data (9/9 revision) we requested broadly confirmed these estimates yielding a reduction in Covid-19 associated with HCQ given within 2 (elapsed time) days of exposure (RR 0.35, 95%CI 0.13 – 0.93; p=0.0438) but not later (RR 0.98, 95%CI 0.67 – 1.45). [8] Recognizing limitations (Supplement) to these estimates, further detail was requested (10/6 revision). Although, the original protocol had intended to enroll only those subjects receiving drug within 4 days from exposure, assuming overnight shipping, 332 and 95 subjects (52% of all subjects) received medication one or two days later than overnight respectively, with 152/821 (19%) subjects receiving drug outside the 4-day window (Table S 3).

### 3.2 Stratification by corrected intervention lag

We stratified subjects according to the corrected intervention lag, consonant with the aim of the original protocol. Mirroring the approach used in Figure S1 of the paper [7] that compares, a priori, the event rates for the two study arms on each day separately, we examined data for each day separately (Table 1). The evident similarities in RR values for numbered days 2-4 justify the pooling of data into “Early” (1-3 days elapsed time) and Late (4-6 days elapsed time, numbered days 5-7) cohorts. We found an HCQ-associated reduction in Covid-19 when received “Early” after exposure from 16.5% to 9.6% (RR 0.58, 95%CI 0.35 – 0.97; p=0.044; NNT 14.5) but not later (“Late”) (RR 1.22, 95%CI 0.72 – 2.04) (Table 1). It is noted that although the event rate for the placebo group at numbered day 4 is higher (18.9%) than for the other days, it is matched by a commensurate change in its respective treatment arm, yielding an RR value (0.624) similar to that for numbered days 2 (0.578) and 3 (0.624). Whether this represents normal variation or has other biological meaning is unclear. Mirroring the later [18] attempt by the original authors to mitigate a similar phenomenon (numbered day 2 for enrollment (Figure S1 in [7]), we compared the event rates in the treatment arm at each numbered day, with the placebo event rate, pooled across all timepoints. While this may obscure a poorly understood relationship between time and development of Covid-19, this approach may be instructive as a sensitivity analysis (Supplement Table S14) which yields the same trends as our primary analysis (Table 1) for reductions in Covid-19 for Early but not Late cohorts. Using 1-3 days elapsed time of intervention lag (numbered days 2-4) for the Early prophylaxis cohort, there is a 32.5% reduction in Covid-19 associated with HCQ (RR 0.674; 95%CI 0.42-1.1, p=0.124). Taking only 1-2 days elapsed time intervention lag, we obtain a 42.9% reduction (RR 0.571, 95%CI 0.30-1.08, p=0.09). This analysis appears to reveal a strong regression line (p=0.033) of Covid-19 reduction and intervention lag (Figure S1).

We performed another sensitivity analysis based on a related follow-up analysis by the original authors [18] using “Days from Last Exposure to Study Drug Start,” a variable previously undescribed in the publication, protocol or dataset. As in a similar PEP study, [19] this variable has limited value because it underestimates, non-uniformly, the intervention lag. Further, this analysis used a modified ITT cohort, rather than the originally reported ITT cohort. [7] Pooling data for days 1-3 and comparing with the pooled placebo cohort yields (Table S 15) a reduction in Covid-19 associated with HCQ on days 1-3 (it is unclear if the “Days from” or “Day on” nomenclature was used) after last exposure from 15.2% to 11.2% (RR 0.74, 95%CI 0.48-1.14, p=0.179). Although not directly comparable, the mITT population used by the authors is somewhat similar to the “Responding Population” which yielded similar results (Table S11).

We did not detect differences in symptom severity scores for either time strata (Supplement). The balance in the demographic and clinical characteristics of the two groups is largely conserved between the Early and Late cohorts (Table 2, Table S 1).

**Table 2:**
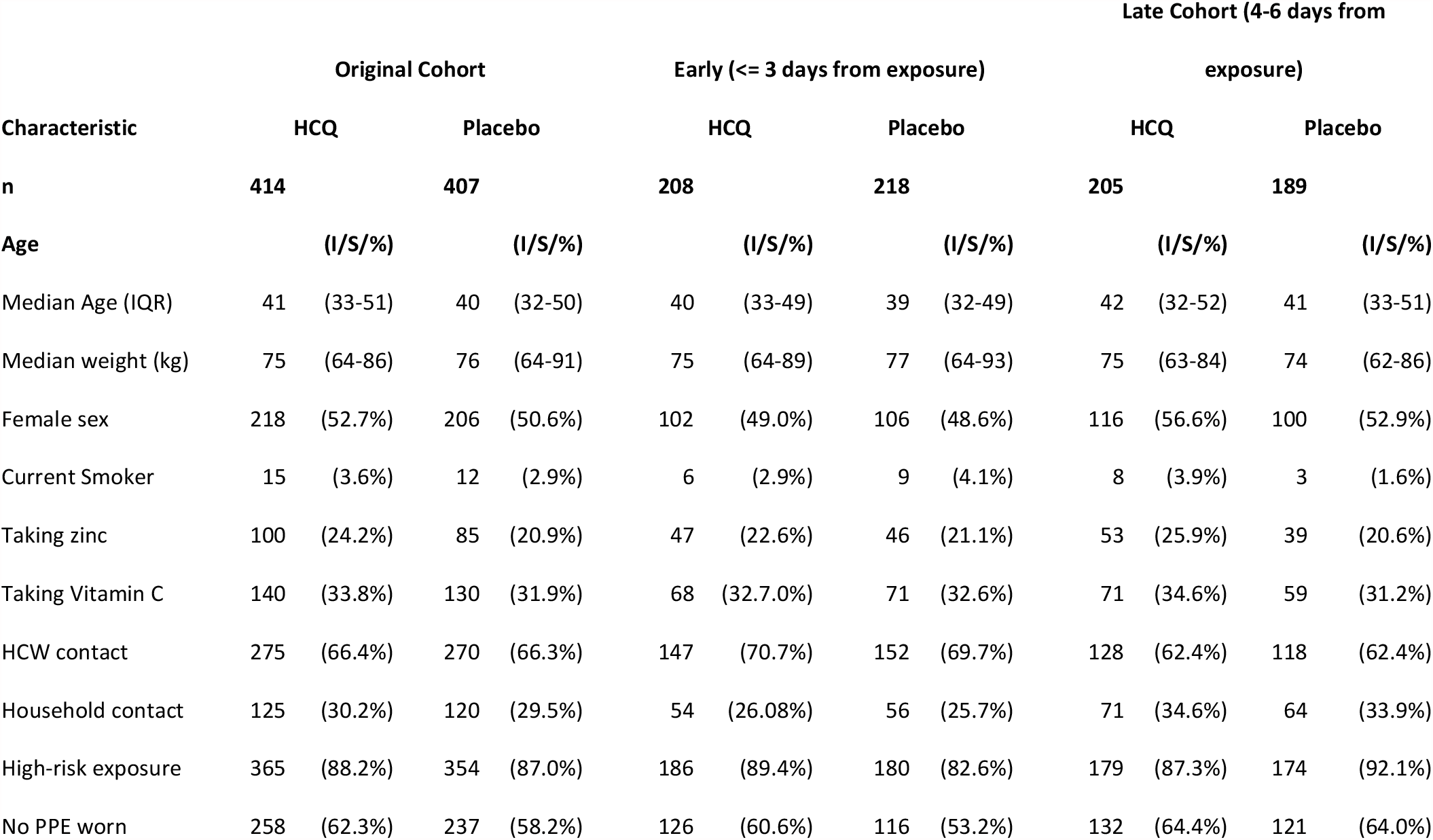
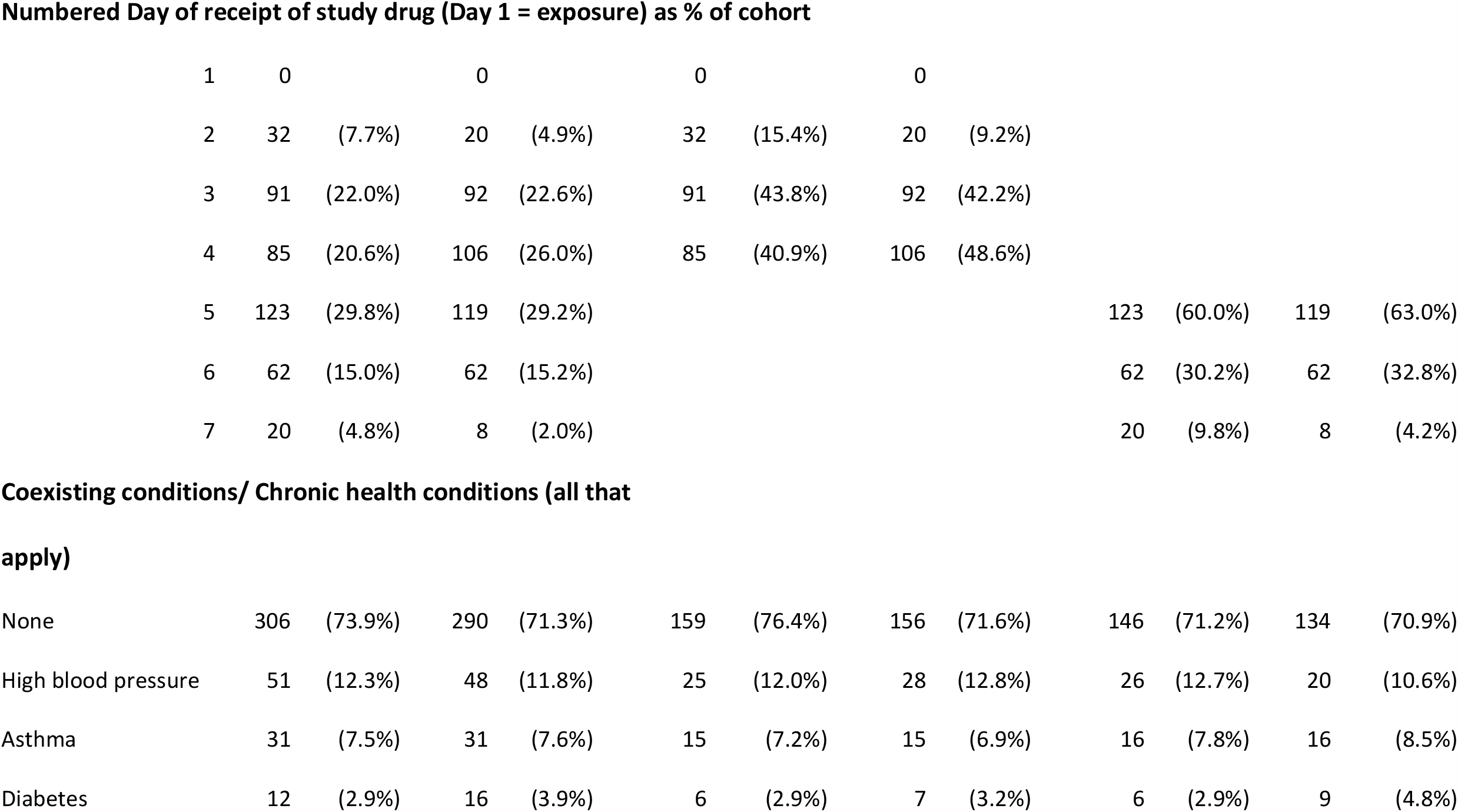
Demographic and clinical characteristics, stratified into Early and Late Cohorts. The data for the original cohort recreates data from the original paper, for comparison and quality control purposes. Several variables have been added. The data are stratified into Early (1-3 days) and Late (4-6 days) post exposure prophylaxis cohorts. See note in Table 1 regarding the nomenclature for “days.” (I/S/%) -Shown in parentheses are interquartile ranges (1^st^ and 3^rd^ quartile), or standard deviations where indicated. All other values within parentheses indicate the percent contribution to the cohort total. See Table S 1 for full list of demographic and clinical characteristics.

### 3.3 Stratification by age, gender

Adopting the same age strata as the PEP study, we found in the Early cohort non statistically significant Risk Ratios of 0.53 (18-35 years), 0.52 (36-50 years), and 2.80 (> 50 years) (Table 3). With no *a priori* reason for selecting these strata, the data are less subjectively supportive of two age strata. Conservatively (Supplement), we set the boundary at 45 years. We found HCQ-associated signal reductions of Covid-19 when given Early in both younger (18-45 years) (RR 0.54, 95%CI 0.29-0.97; p=0.0448, NNT 11.5) and older (>45 years) (RR 0.75, 95%CI 0-27-2.05) subjects. Within the Early and Late cohorts, gender-dependent effects were not apparent (Table S 4).

**Table 3:**
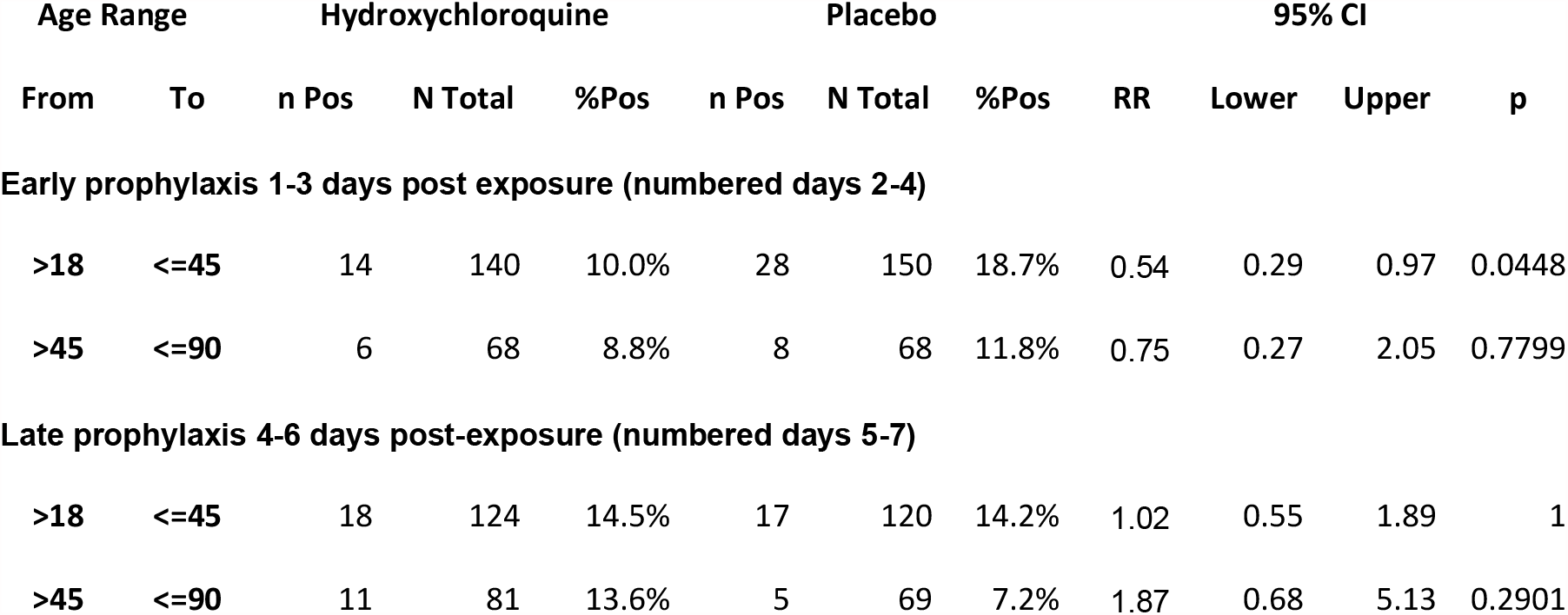
Stratification of effect associated with hydroxychloroquine by age based on time from exposure to drug receipt (ITT population) The number (and percent) of subjects with a Covid-19 positive outcome are shown for each group along with the total number of subjects for that group, stratified by time from exposure to drug receipt. The elapsed time range in days is shown for Early and Late cohorts. See note in Table 1 regarding the nomenclature for “days.”

| Age Range |  | Hydroxychloroquine |  |  | Placebo |  |  | 95% CI |  |  |  |
| --- | --- | --- | --- | --- | --- | --- | --- | --- | --- | --- | --- |
| From | To | n Pos | N Total | %Pos | n Pos | N Total | %Pos | RR | Lower | Upper | p |
| **Early prophylaxis 1-3 days post exposure (numbered days 2-4)** |  |  |  |  |  |  |  |  |  |  |  |
| >18 | <=45 | 14 | 140 | 10.0% | 28 | 150 | 18.7% | 0.54 | 0.29 | 0.97 | 0.0448 |
| >45 | <=90 | 6 | 68 | 8.8% | 8 | 68 | 11.8% | 0.75 | 0.27 | 2.05 | 0.7799 |
| **Late prophylaxis 4-6 days post-exposure (numbered days 5-7)** |  |  |  |  |  |  |  |  |  |  |  |
| >18 | <=45 | 18 | 124 | 14.5% | 17 | 120 | 14.2% | 1.02 | 0.55 | 1.89 | 1 |
| >45 | <=90 | 11 | 81 | 13.6% | 5 | 69 | 7.2% | 1.87 | 0.68 | 5.13 | 0.2901 |

### 3.4 tratification by co-morbidities and contact type

Considering only subjects reporting no co-morbidities (particularly excluding asthma and co-morbidities classified as “other”), suggested stronger HCQ-associated effects within time-or age-related strata (Supplement). Differences in HCQ-associated effects of Early prophylaxis between Household (RR 0.69) and HCW (0.92) contacts reached statistical significance after time-stratification with a reduction of Covid-19 in household contacts (RR 0.35, 95%CI

0.13-0.89; p=0.025, NNT 5.7, Table 4). Differences in the baseline incidence of Covid-19 and the relationship between contact type, risk level and changes in risk definitions are described in the Supplement.

**Table 4:**
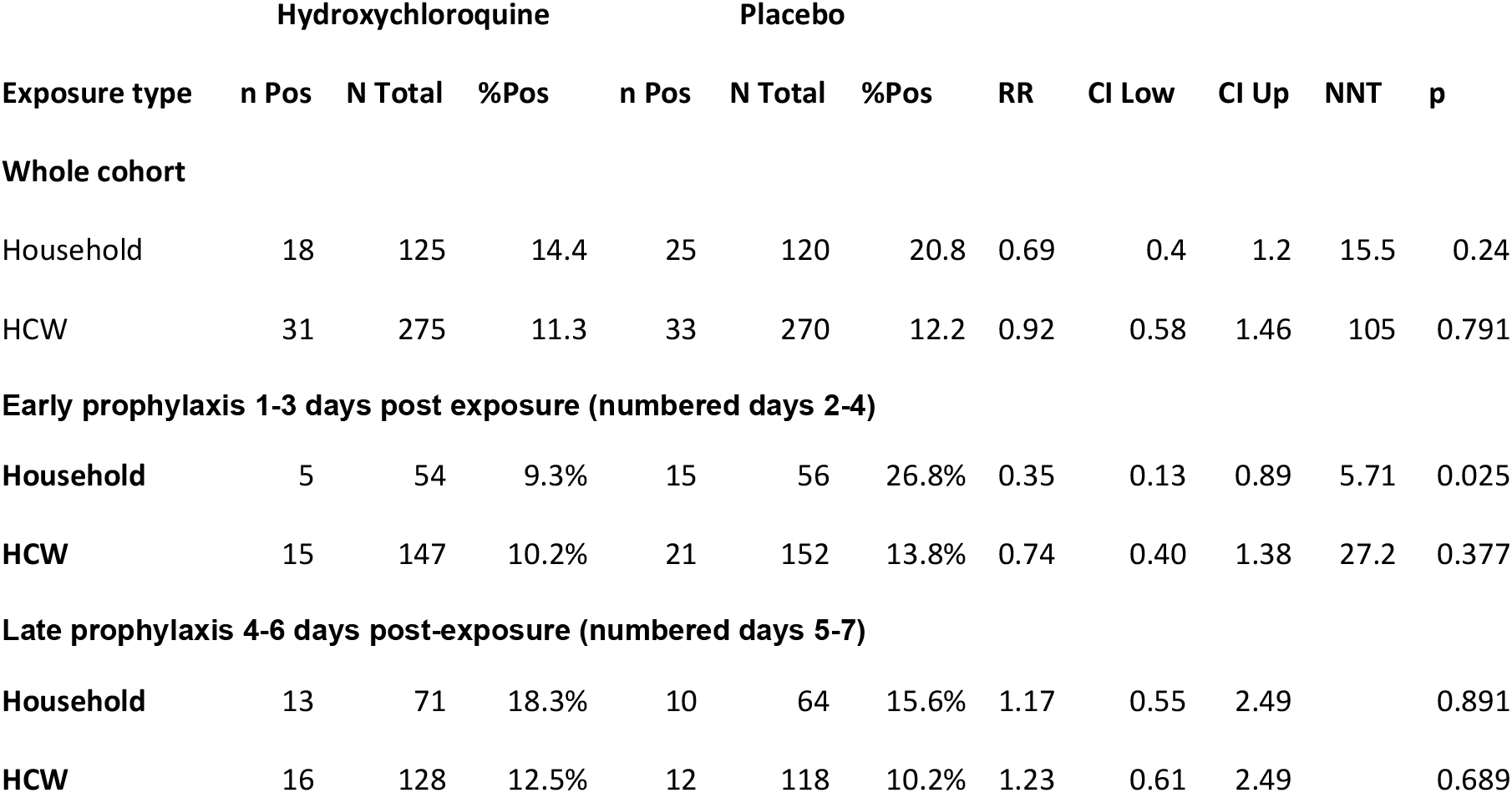
Stratification of effect associated with hydroxychloroquine by exposure type based on time from exposure to drug receipt (ITT population) The number (and percent) of subjects with a Covid-19 positive outcome are shown for each group along with the total number of subjects for that group, stratified by time from exposure to drug receipt. The elapsed time range in days is shown for Early and Late cohorts. See note in Table 1 regarding the nomenclature for “days.”

### 3.5 Adherence to protocol, use of folate placebo, ex-protocol use of zinc and ascorbate

Similar trends were obtained after excluding subjects not contributing outcome data or taking study medication. Stratifying into Early and Late prophylaxis cohorts revealed no discernible folate-associated effect (Supplement). With poorly detailed observational data, there did not appear to be zinc-or ascorbate-associated effects (Supplement). The use of zinc and ascorbate appears balanced between the groups in the whole and Early and Late time cohorts (Table 1).

## 4 Discussion

### 4.1 Overall Findings

In tackling our primary objective of defining temporal effects of HCQ, we understood that HCQ prophylaxis had been “*initiated within 4 days after* […] *exposure*.” [7] Although others, [11],[13],[14] including the authors of NIH [20] guidelines and the editorial [5] accompanying the paper, shared this understanding, it requires revision for two main reasons.

Firstly, after obtaining missing data, we found that many participants received medication after the intended overnight delivery or after four days from exposure, independently conjectured. [21] A similar issue pertains to the companion PET study. [15] Secondly, inconsistent terminology (“Day on” vs. “Days from”) led to overestimating the time from exposure to enrollment or to drug receipt by one day. Correcting these understandings yields a 42% statistically significant reduction of Covid-19 associated with HCQ received between 1 and 3 days (elapsed time) after exposure, but not later. To mitigate the effects of possible outlying values in the placebo event rate, we used in a sensitivity analysis, the same, albeit limited, approach employed to a related problem by the original authors in a follow up work [18]. Another sensitivity analysis based on this follow-up work yielded a similar trend based on the time from last (rather than highest risk) exposure, a parameter of limited value not previously used or released by the original authors. Both approaches yielded support for our overall conclusions suggesting that early intervention with HCQ is an effective strategy. The early use of HCQ is supported by estimates for an incubation period of 3-8 days. [22]

We found an age-dependent, statistically significant reduction of Covid-19 associated with Early prophylaxis. Re-analyzing the same PEP dataset without time stratification, we confirmed (Supplement) Luco’s [13] report of HCQ-associated reductions in Covid-19 in subjects younger than 50 years, reaching statistical significance in the high-risk exposure cohort. Interpretation of age-related effects is limited by a poorly understood relationship between age and Covid-19 susceptibility, increases in incubation period with age [23] and a low representation of older subjects with a low baseline incidence of Covid-19.

Small co-morbidity subgroups prompt cautious interpretation. However, co-morbidities attenuated age-and time-dependent HCQ-associated effects. Although age-related responses associated with HCQ may be related to co-morbidity, [13] excluding co-morbid patients did not yield equivalent effects in age strata. Asthma and “other” co-morbidities contributed most to attenuating the HCQ-associated response, consistent with an independent re-analysis. [13]

Mirroring the original data, we found a significant HCQ-associated effect in household contacts. This result may reflect differences in access to advanced PPE, hygiene training, likely multiple exposures, and the ability to quarantine after exposure. Thus, household contacts in this study may share much with first responders in the companion PrEP study, [24] where a 64% HCQ-associated reduction in Covid-19 was observed (combined dose groups). Further, the changing risk definitions and their apparently inconsistent application between contact types may confound understanding of how contact type and risk level affect Covid-19 development. Whether the apparent lack of an HCQ-associated effect in moderate-risk exposures is a statistical aberration or is biologically meaningful is unclear.

With a small “no folate” cohort, we did not detect an effect of folate, but cannot exclude a confounding effect, suggested by a growing literature on the subject [25] (Supplement). Certainly, using folate as a placebo does not seem prudent. Due to a paucity of data, we could not determine whether there was an effect of zinc or ascorbate other than noting no differences associated with HCQ in subjects taking neither agent and the entire ITT cohort.

### 4.2 Findings in the context of other Covid-19 studies

Our findings are made in the climate of concern [26] for the reliability of publications related to Covid-19 and the controversy surrounding HCQ. [27],[28] This is partly fueled by a widening understanding of Covid-19 pathogenesis and the multiple, sometimes stage-dependent, mechanisms proposed for HCQ. [29]

In hospitalized patients, factors confounding HCQ’s effectiveness are beginning to be better understood [30], with clear reports of significant HCQ-associated reductions in mortality with the use of zinc. [3],[31] The possibly synergistic use of steroids [32] may further confound some studies. At earlier stages, any HCQ-associated effect appears independent of zinc, evinced (weakly) by the lack of synergy we observed. Further, using zinc may be futile in otherwise healthy, especially younger subjects with no zinc deficiency or dysregulation.

### 4.3 Findings in the context of other prophylaxis studies

For prophylaxis, understanding differences in age, gender, quarantine practices, testing methods, co-morbidities, and the possibility of multiple “index” exposures, appear important in reconciling apparently conflicting studies (Table 5). Particularly important are differences in intervention lag which may be extended beyond an effective range due to HCQ’s long half-life and the time taken to reach steady state. Watanabe’s [14] confirmed (Table S 7) estimate illustrates this. After censoring Covid-19 events occurring before completion of five days of treatment, there is greater HCQ-associated effectiveness in the whole population (RR 0.59, 95%CI 0.32-1.07, p=0.085) and to some extent in the Late prophylaxis cohort.

**Table 5:**
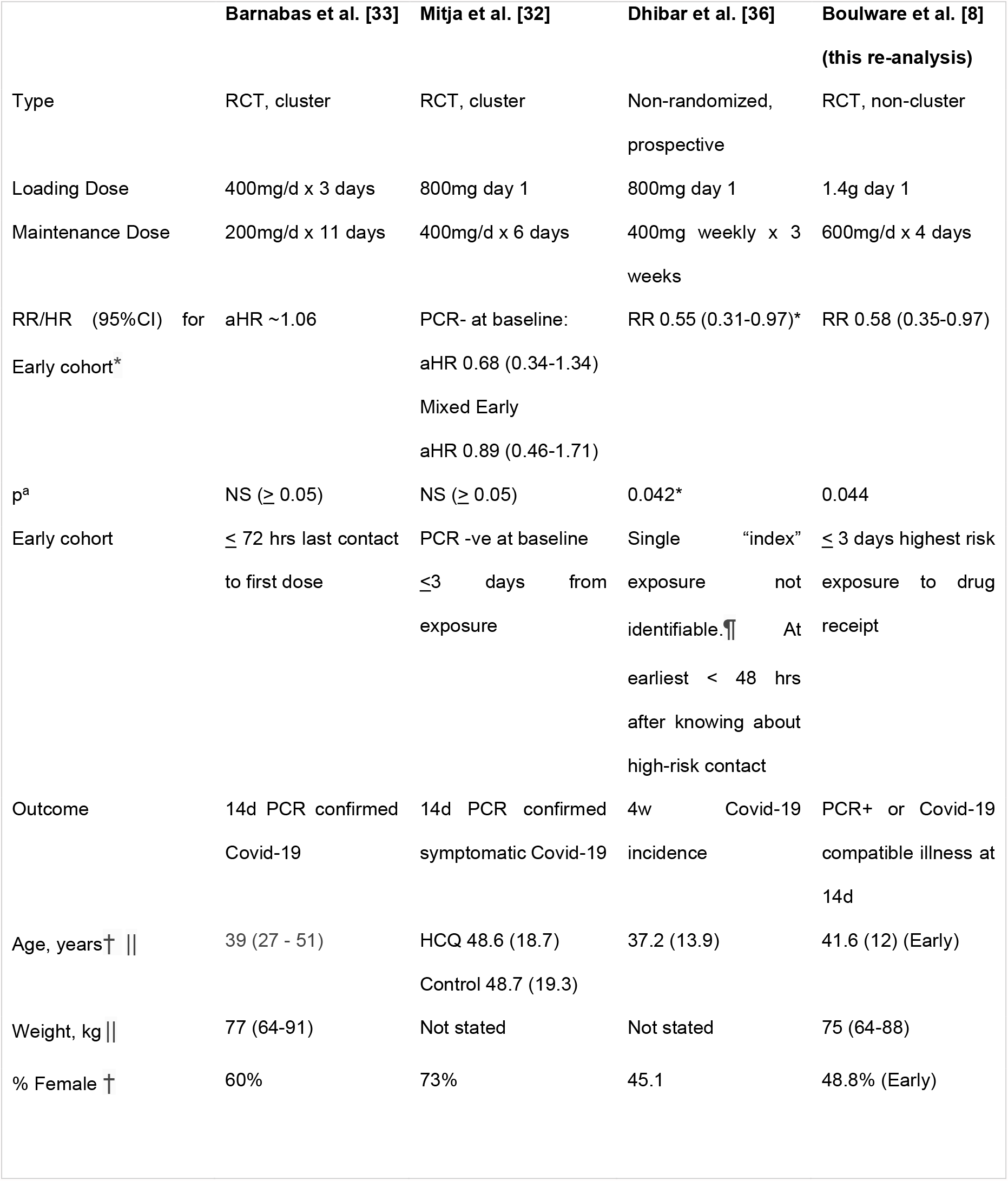
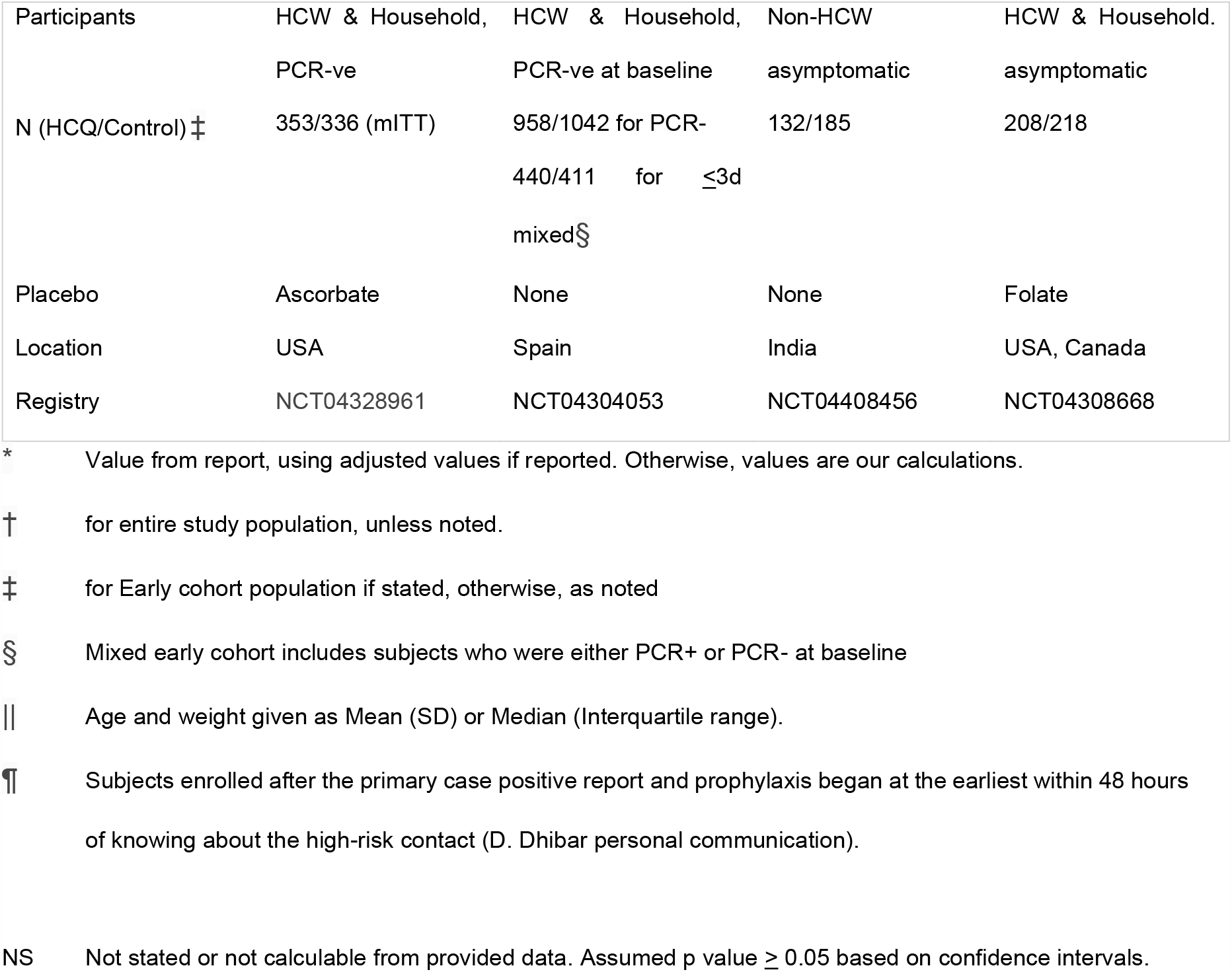
Comparison of HCQ-associated effects after early prophylaxis in similar PEP studies.

These differences may be amplified by inadequate loading and maintenance doses, contributing significantly to differences between outcomes of the PEP and two similar studies (Mitja et al., [33] Barnabas et al. [19]). This is modeled in pharmacokinetic simulations [34] conducted to support the PEP study. Although qualitatively useful, the model is limited [24],[34] and does not consider other factors such as age, obesity and genetic influences.

In a Spanish cluster-randomized trial (Mitja et al., [33]) more modest HCQ-associated effects were seen with loading and maintenance dosing that were more modest than in the PEP study (Table 5). Although there were poor HCQ-associated effects with a short (<= 3 days) intervention lag (RR 0.89), there were differences according to PCR test status at baseline. For PCR-negative patients there was an effect signal (aRR 0.68, 95%CI 0.34–1.34), but not for PCR-positive subjects (aRR 1.02; 95%CI 0.64–1.63). Becoming PCR positive describes the moment beyond which a drug is unlikely to be effective possibly more reliably than a particular intervention lag. Accordingly, this study supports our findings suggesting a beneficial effect of early intervention. Possibly also attenuating HCQ-associated effects is a mean age (48.6 years) higher than in the PEP study (41.6 years).

No HCQ-associated effect was reported in a household-randomized (Barnabas et al. [19]) study (USA) that used the lowest loading and maintenance dosing among comparable studies (Table 5), speculated by the authors to have been insufficient, and likely ineffective according to the PEP study’s pharmacokinetic simulation. [34] By day 14, no reduction in infection rate, determined by PCR testing of self-collected nasal swabs, was associated with HCQ even for intervention lags under 72 hrs. The substantive lack of a loading dose may have extended the intervention lag beyond an effective range. Non-uniform underestimation of intervention lag based on the time from last rather than first contact, likely obscured any time-dependent effect. The possibly anti-viral effect of the ascorbate [35] placebo may have confounded the results. Although data were not age-stratified, the population was of a similar age to the PEP study, and with a higher representation of Hispanic subjects.

Illustrative of the limitations of the pharmacokinetic model [34] is an Indian non-randomized prospective study. [36] Despite using lower loading and maintenance doses than in the PEP study (Table 5), this study reported a 45% reduction of Covid-19 associated with HCQ at 4 weeks.

Differences in HCQ-associated responses between “Early” and “Late” PEP illustrate the poorly defined position that PEP occupies on a continuum between PrEP and PET. The variable possibility of other exposures occurring before and after a single “index” exposure means that the Early PEP cohort has much in common with the population of the companion PrEP study. [24] Although the PrEP study was hampered by poor recruitment, once or twice weekly use of HCQ (after a loading dose) in HCW was associated with reduced development of Covid-19 by 27%, compared with folate placebo (HR 0.73, CI 0.48-1.09, p=0.12, combined groups). It is unclear why a thrice-weekly schedule, as advocated by the authors’ own pharmacokinetic model, [34] was not used. Age related HCQ-associated effects were of a similar order of magnitude (34-45%) in the PEP and PrEP studies (Supplement Table S 6).

### 4.4 Limitations

Limitations related to post hoc and subgroup analysis [37] are partially offset by our use of two-sided tests, when directionality in the original data may justify otherwise. It is noted that due to its early termination, the cohort of the original study represents a subgroup subject to similar interpretative limitations. Given the enormity of the pandemic, the high cost of falsely rejecting beneficial drugs with Type II errors related to underpowering and early termination could justify greater toleration for Type I errors and other statistical challenges. Methods have been proposed to consider the relative costs of incurring Type I or II errors in calculating appropriate significance levels. [38] Our primary time stratification based on newly-acquired data essentially represents the *a priori* analysis intended by the original authors. Nonetheless, this remains a *post hoc* study; results should be interpreted accordingly, and hypotheses tested in prospective studies sufficiently powered to accommodate multiple comparisons in sub-groups.

Our analysis retains the limitations acknowledged by the original authors related to the availability and access to testing, the use of a clinical definition of Covid-19, the reliance on self-reported data and the generally young population studied. There are other limitations. The study poorly represents minority, African-American, Hispanic and Latino populations. The rapidly executed study overcame logistical challenges to rapidly collect real-world data, with advantages and disadvantages of a pragmatic design. [9],[39] Self-selection bias inherent in this type of study may have been compounded by FDA cautions regarding HCQ. [4] Unlike similar studies, [19],[33] the PEP study was not cluster-randomized. [39]

Other limitations relate to the estimation of the interval between exposure and treatment with 24-hour windows of uncertainty on either side. The earlier window is due to subjects providing only the date of their highest risk exposure. The later window is due to de-identification of shipping data, and the unknown interval before ingesting the first dose. The original authors (personal communication) attempted to minimize this by delivering medication to where the participant knew they would be at its expected arrival time.

Time-related or other biases may be associated with the exclusion of 100 randomized subjects who became symptomatic before medication was received and aggregated into the companion treatment study. [15] Lastly, analysis of the effect of risk level is confounded by an inability to discriminate between nuances within the risk categories.

## 5 Conclusions

Analysis [40] of the PEP, [7] companion [15],[24] and other [19],[33] studies raise no significant safety concerns for using HCQ in the populations studied. Integrating a public dataset with novel unpublished data concerning unaccounted-for shipping times, we found that, especially in younger subjects, hydroxychloroquine was associated with significantly reduced illness compatible with Covid-19 when initiated between 1 and 3 days (elapsed time) after a high-risk or moderate-risk exposure at higher loading and maintenance doses than in similar studies.

Combined with other post hoc analyses, our findings controvert the conclusions of the original study, warranting prospective confirmation.

## Supporting information

IJME - COI Disclosure

## Data Availability

Microsoft Excel files will be available on reasonable request up to one year after publication to qualified investigators subject to an agreed upon data sharing agreement. The source dataset is available from the original authors.

## Abbreviations

HCQ: Hydroxychloroquine (and its salts)
HCW: Health Care Worker
ITT: Intent to Treat
mITT: Modified Intent to Treat
NNT: Number Needed to Treat
PCR: Polymerase Chain Reaction
PEP: Post-Exposure Prophylaxis
PET: Post Exposure Treatment
PPE: Personal Protective Equipment
PrEP: Pre-Exposure prophylaxis
RCT: Randomized Clinical Trial

## Acknowledgements

We thank the original authors for clarifications provided, collecting additional shipping-related data at our request and providing confirmatory calculations. We also thank Drs. Marcio Watanabe, Juan Luco and Philip Lavin for their valuable comments. This acknowledgment does not imply endorsement of our work.

## Funding

This research did not receive any specific grant from funding agencies in the public, commercial, or not-for-profit sectors.

## Conflicts of Interest

The sponsor is entirely responsible for its design and conduct. The sponsor and principal investigator have no financial or other conflicts of interest in the subject matter of this study. DMW is the president of Synechion, Inc. and KevMed, LLC, providing services for the medical industry and marketing medical products, respectively, outside of the area of this work. See ICMJE forms for further details. DM is the president of ZSX Medical, LLC. developing surgical devices and a Principal at Third Eye Associates, a technical consulting company. PK and SAS report no conflicts.

## Notes

### Competing Interest Statement

The authors have declared no competing interest.

### Clinical Trial

NCT04308668

### Clinical Protocols

https://osf.io/hyp8k/

https://www.medrxiv.org/content/10.1101/2020.08.19.20178376v2

### Funding Statement

There is no external support for this study.

### Author Declarations

No IRB approval was required for this work. This is a re-analysis of a de-identified, publicly released dataset obtained from an IRB-supervised study. Additional requested data were de-identified data and provided after we had been advised that the Privacy Officer of the University of Minnesota would be consulted to ensure regulatory compliance.

### Summary of Updates

Versions 3.1-3.4: Revisions since version 2 of this manuscript was pre-printed December 12, 2020. i)Analysis confirming Watanabe, considering subjects becoming Covid-19 positive after the five-day treatment period (Table S 7). Comment added in discussion. ii)Abella, Bhattacharya. references and comments removed (to preserve wound count and reference limit) iii)Yang [37] reference inserted, noting their independent observation regarding the shipping time issue, recently brought to our attention. iv)Revised discussion of statistical limitations, Mudge et al., reference added. v)Plain language summary moved to Supplement vi)CDC reference [38] removed, duplicates information in WHO reference. [39] vii)Pharmacokinetic study by Boulware group referenced [40] and integrated into discussion of comparable studies. viii)Pragmatic trial reference replaced with better citation. ix)Table 5, weight added x)Typos and word economy, update references and format xi)Running and extended title xii)Clarifications and additional analyses to address comments made on medrxiv. These do not affect study results or interpretation. a.Clarification regarding <day on> vs. <days from> nomenclature added throughout. b.Clarification of variables used in different versions of the dataset (Supplement), including a newly circulating, but inaccurate revision (10/26). c.Sensitivity analysis using pooled placebo cohort to compare time-related effects. d.Sensitivity analysis based on Figure 1 of Nicol et al. [26] xiii)Title change to <novel, missing, data> xiv)Highlights added

