## Supplemental Appendix for "Effective post-exposure prophylaxis of Covid-19 associated with hydroxychloroquine: Prospective dataset re-analysis incorporating novel, missing data"

Running Title: Postexposure hydroxychloroquine Covid-19 prophylaxis: Reanalysis, novel data

David M. Wiseman, PhD, MRPharmS; Pierre Kory, MD, MPA; Samir A Saidi, PhD, MB ChB; Dan Mazzucco, PhD.

### Authorship Contributions

DMW conceived of the study with additional input from PK and DM. Statistical analysis was performed by DMW and DM. DMW wrote the first draft of the manuscript and is overall study guarantor with contributions from PK, DM and SAS. SAS contributed to analysis. All authors reviewed, revised, and approved the manuscript.

**Version and Word Count**
Version 3.4 07/02/21
*Main text*
Abstract 249 words
Text 3981 words
40 References
5 Tables

*Supplement*
15 Tables
1 Figure
40 References
Plain language summary 418

**Abbreviations**

| | |
|---|---|
| 95%CI | 95% Confidence Intervals |
| aHR | adjusted Hazard Ratio |
| aRR | adjusted Risk Ratio |
| CDC | Centers for Disease Control |
| CQ | Chloroquine (and it salts) |
| EUA | Emergency Use Authorization |
| FDA | United States Food and Drug Administration |
| HCQ | Hydroxychloroquine (and its salts) |
| HCW | Health Care Worker |
| HR | Hazard Ratio |
| IRB | Institutional Review Board |
| ITT | Intent to Treat |
| LTF | Lost to Follow Up |
| NIH | National Institutes of Health |
| NNT | Number Needed to Treat |
| NO | Nitric oxide |
| OR | Odds Ratio |
| OSF | Open Science Framework (https://osf.io/) |
| PCR | Polymerase Chain Reaction |
| PEP | Post-Exposure Prophylaxis |
| PET | Post Exposure Treatment |
| PPE | Personal Protective Equipment |
| PrEP | Pre-Exposure prophylaxis |
| RCT | Randomized Clinical Trial |
| RP | Responding Population |
| RR | Risk Ratio |
| SOC | Standard of Care |

**Plain Language Summary**
A highly-cited clinical trial examined the ability of hydroxychloroquine (HCQ) to prevent Covid-19 just after an exposure to a person confirmed to have Covid-19. There was an HCQ-associated reduction of Covid-19 by an overall 17%; 36% in younger subjects, and 49% in subjects given HCQ very early after exposure. Likely because the study had too few patients to find what may have been a medically and economically meaningful, reduction, this effect was not statistically significant.

Studying the trial data, we discovered an unintended and variable delay in the delivery of study drug which may have masked any drug effect. The investigators provided further information missing from the original analysis at our request that confirmed our theory. About half of the participants received drugs one or two days later than intended, about a fifth beyond the four days the investigators thought the drug might work.

When we factored in this new information, we found that if HCQ was given early (up to three days after exposure), it was associated with a statistically significant 42% reduction of Covid-19. Giving HCQ later had no effect. There was a greater effect in younger (less than 45 years) rather than older subjects (47% vs. 25%). Gender did not seem to affect the results, but there was a greater HCQ-associated reduction (65%) when it was given early to people exposed to Covid-19 in a household environment rather than to health care workers (26%). The effects associated with HCQ were better in people without co-existing conditions.

These re-calculations are important because the study, as originally analyzed, was the only randomized study that dealt with preventing Covid-19 cited by FDA to support a key public health decision made in June 2020 regarding HCQ. It is only one of only three studies for this sort of prevention cited by the Treatment Guidelines of the National Institutes of Health. Although other studies have shown that the drug is not effective to treat established cases of Covid-19, our research suggests that that it is effective for prevention. Other prevention studies have failed to show a benefit of HCQ, possibly because they have used lower doses or have estimated the timing of dosing differently. Our research paves the way for our result to be confirmed under clinical trial conditions and for a re-examination of public health policy regarding this drug. Even with vaccination, there remains a need for approaches like this to prevent Covid-19 while individual and community immunity develops, and in preparedness for possible vaccine inefficacy against new variants.

**Short Summary**
A prospective re-analysis of a public dataset integrated with novel data found an HCQ-associated reduction of illness compatible with Covid-19 when received between 1 and 3 days after a high-risk or moderate-risk exposure (RR 0.58, 95% CI 0.35-0.97, p=0.044, NNT14.5).

**Supplemental Background**
*Folate, Zinc and Covid-19*
Possibly confounding the study is the use of folate for the placebo for which a number of actions have been noted or speculated in Covid-19. [1]. In silico, folate may interact with SARS-Cov-2 [2],[3] as well as with furin. [4] Blood folic acid levels were significantly lower in severe Covid-19 patients. [5] There may be an association between folate and severity or prevention of disease with other viruses. [6],[7],[8],[9] Modulation of endothelial function may be a fruitful approach in Covid-19 by improving pulmonary perfusion and reducing hypoxemia. High doses of folic acid (with Vitamin B6) improve NO mediated vasodilation in diabetic children [10] at doses of a similar order of magnitude (5mg) as those used in the PEP study (2.8mg initially, then 1.2mg daily, confirmed by Boulware et al., personal communication). Folate has been proposed to be a protective factor for Covid-19 in pregnant women. [11] Since immunogenicity to a mRNA COVID-19 vaccine, is hampered by methotrexate a “folate antagonist”, this may indicate a positive effect of folate on the immune response to Covid-19. [12]

Conversely, there may be a negative effect of folate in Covid-19. The folate receptor on macrophages is upregulated in inflammation. [13] *In vitro* inhibition of viral replication by methotrexate [14],[15] is synergistic with remdesivir and rescued by folinic acid. [14] Folate deficiency inhibits the proliferation of CD8+ T cells in vitro. [16] Methotrexate may be useful in Covid-19 [17] (with folinic acid rescue [18],[19]) possibly via effects on lymphocytes. [20]

Given the much-discussed effect of zinc as a treatment for viral infections in general and Covid-19 in particular, [21],[22] and the action of CQ as a zinc ionophore, [23] the ex-protocol use of zinc supplements may have confounded the data. A similar case can be made for the ex-protocol use of Vitamin C. [24]

**Supplemental Methods**

*Protocol History*

Our protocol was registered on August 13, 2020 (v1.0, osf.io/fgd53/) and revised August 19, 2020 (v1.1, osf.io/9rpyt) before accessing the initial PEP dataset. We issued version v1.2 (September 27, 2020, osf.io/vz8a7/ [25]) prior to receiving additional data regarding the time to drug receipt in the 10/6 revision.

*Clarifications to Dataset, Quality Control*

In reviewing the PEP study dataset, we verified variable tallies with the published account. After notifying the original authors, most of the discrepancies were resolved in the 9/9 revision. We performed similar checks on the 9/9, 10/6, and 10/30 revisions. Several of the original analyses were recreated to verify data importation and processing. A record of our questions and clarifications received was appended to our protocol registration (osf.io/udx28/).

There were four main areas requiring clarification (see Supplemental Methods) related to (i) tallies and definition of high- and moderate- risk exposure; (ii) tallies for subjects adhering to study medication; (iii) estimation of time from exposure to receipt of study medication ("intervention lag"); and (iv) nomenclature describing timing of study events.

Firstly, from the various combinations of reported use of different PPE items, we were unable to recreate the tallies for the number of subjects having high or moderate risk exposures. We were informed that the published risk definitions had changed over time, and that an erratum would be submitted (Table S *2*).

Secondly, we could not recreate the tallies for subjects adhering fully, partially or not at all to the study medication. The authors provided a variable (9/9 revision) from which, combined with other variables, we could recreate these tallies.

Thirdly, the published data had been stratified according to the days from the reported date of highest risk exposure to enrollment, rather than the time from exposure to first dose. Requesting further detail, additional data were provided (9/9 revision) describing the time from enrollment to receipt of study drug. These data did not account for time zone differences, delivery times for Canadian participants or biases introduced because of estimates of the time from exposure to enrollment. At our request, the authors provided (10/6 revision) the number of days from exposure to receipt of drug, likely self-correcting for time zone differences. This included data for the Canadian subjects.

Fourthly, the authors notified us of a third dataset (10/30) revision clarifying the previously used nomenclature describing the timing of study events. The word "Day" was clarified to represent the numbered day on ("Day on") which enrollment (Days 1-4) or study drug receipt (Days 1-7) occurred, with the date of highest reported exposure being "Day 1." As clarified, this terminology is used, respectively, in the "exposure_days_to_enroll: and "exposure_days_to_drugstart" variables provided by the authors.

Accordingly, to calculate the elapsed time between exposure and enrollment or receipt of study drug ("Days from"), one day must be subtracted from the stated numbered "Day." In the 10/30 revision, the definition for the variable describing the time from exposure to enrollment was partially clarified, but not that describing time from exposure to receipt of study drug. A derivative version of this variable ("Exposure_to_DrugStart") was provided whose individual values were all smaller than for its primary version ("exposure_days_to_drugstart") by one day. To further confuse matters, we are aware of a clearly inaccurate 10/26 version (filename PEP_Public_Data_01Oct2020.csv, timestamp 10/26/2020 5:36pm) of the dataset that has been provided to some colleagues as recently as May 2021 in which the values contained in these two variables are incorrectly identical, rather than correctly differing by one day as reported in the 10/30 version.

Adopting this clarification, we note its inconsistency with statements made in the original paper and a subsequent work [26] that indicated the occurrence of some study events to be one day later. This clarification does not alter the relative time stratification we present here. We have endeavored in this paper to distinguish between the use of the “Day on” and “Days from” nomenclature.

*Provision of outcome data, adherence to study drug, use of folate and comorbidities*
We also analyzed the data according to adherence to taking study medication, whether subjects provided outcome data and the use of the folate placebo. We constructed a “Responding Population” (RP) by excluding those subjects who were lost to follow up (LTF) or withdrew consent and who provided no outcome data (Table S 8).

To examine the effect of folate and adherence to study drug, we constructed three treatment arms. In addition to the HCQ treatment arm (fully and partially adherent) we constructed a “folate only” control arm (fully or partially adherent) and a “no folate” control arm consisting of the Canadian subjects randomized to lactose placebo pooled with subjects identified as taking neither HCQ or placebo. Within the ITT population, we examined the effect of co-morbidities on any possible effect of HCQ. Due to the low incidence of most of the co-existing conditions, we examined only the three most frequent conditions (asthma, diabetes, hypertension), in addition to subjects reporting no co-existing conditions or conditions not otherwise listed in the screening questionnaire.

*Confirmation of findings by Luco*
We performed several analyses to replicate the findings of Luco [27] who conducted his own re-analysis of the same PEP dataset, related to the effect of age, exposure risk and co-morbidities. These analyses also serve as a verification of some of our calculations.

*Confirmation of Findings by Watanabe*
We performed an analysis to confirm the finding of Watanabe [28] concerning the consideration of only those subjects who developed a primary outcome later than five days, the period of the full course of treatment with drug.

*Comparison with findings of Rajasingham et al. in PrEP study*
The age strata used in the pre-printed companion PrEP study [29] (see Revision History below) were revised in the published version [30] permitting comparison of age-related findings for the PrEP and PEP studies.

**Supplemental Results**
*Verification of primary time stratification*
As a further quality check, we verified our primary time-stratification with two calculations provided by the authors of the original study. These calculations used the new data we had requested describing the time between exposure and receipt of study drug provided in the 10/6 revision.

As provided, both calculations appear to refer to elapsed time (“Days from”), but as later clarified actually refer to the numbered day (“Day on”) nomenclature, i.e. subjects receiving study drug on numbered Day 3 or earlier, with Day 1 = day of exposure.

The first calculation provided was: *“By ITT analysis for those <=3 days from exposure to med delivery, the actual Odds Ratio = 0.53 (95%CI, 0.23 to 1.22, P=0.14).”* With likely minor differences due to rounding, we replicated this calculation with OR=0.5310 (95%CI 0.2302-1.2246; p=0.149).

The second calculation provided was: *“By modified intent-to-treat (limiting to those who took a dose of the study drug within <=3 days of exposure), Odds Ratio = 0.57 (95%CI, 0.24 to 1.34, P=0.20)”*. With likely minor differences due to rounding, we again replicated this calculation (OR 0.5704, 95%CI 0.2433-1.3375; p=0.2073).

*Effect of exposure risk on outcomes*
There were differences in the response associated with HCQ noted according to the level of risk exposure (Table S 5 - A), with responses in the high-risk group numerically better (RR 0.77) than in the moderate-risk group (RR 1.66). These differences reached statistical significance after time-stratification for Early HCQ prophylaxis in subjects experiencing high (RR 0.48, 95%CI 0.28-0.85, p=0.013, NNT 10.9) but not moderate (RR 1.73, 95%CI 0.48-6.23) risk exposures. The population size for the moderate risk level was small, especially for the Late prophylaxis cohort.

Numerical differences in HCQ-associated effects in the whole cohort between Household (RR 0.69) and HCW (0.92) contacts reached statistical significance after time-stratification with a reduction of Covid-19 in household contacts after Early HCQ prophylaxis, compared with placebo (RR 0.35, 95%CI 0.13-0.89; p=0.025, NNT 5.7) (Main body, Table 4).

A higher baseline incidence of Covid-19 was noted in Household contacts than for HCW, the largest difference being seen in the Early prophylaxis cohort (26.8% vs. 13.8%). The lower baseline incidence in HCW may partly be related to a higher complement of moderate-risk exposures (~14%) than in Household contacts (~6%) (Table S 5 B). Further, since the exposure risk definitions had been revised (Table S 2), what was considered "moderate risk" for a Household contact, may have been classed as high risk for a HCW. Accordingly, the HCW cohort may have had an even higher proportion of moderate-risk exposures.

*Age-dependence of HCQ-associated effect, choice of strata*
Adopting the same age strata as the PEP study, we found in the Early cohort non statistically significant Risk Ratios of 0.53 (18-35 years), 0.52 (36-50 years), and 2.80 (> 50 years). With no a priori reason or justification for selecting these strata, the data are less subjectively supportive of two age strata. The RR (0.52, 95%CI 0.31-0.9, p=0.016, NNT = 10.6) for the combined lower age (18-50) strata has the lowest (0.9) upper 95%CI of all strata extending from 18 years upwards. This, however, leaves the older age stratum with only one and three Covid-19 positive events in the control and HCQ groups respectively, impairing any interpretation of age-related effects. Accordingly, on a post hoc basis we set the boundary at 45 years where the upper 95%CI for the lower stratum was still <1, and where the upper stratum retained higher numbers of Covid-19 events in the two groups (HCQ, 5; placebo, 8). Several methods [31] have been proposed to define strata which we attempted to accomplish empirically on an exploratory basis.

We verified the findings of Luco [27] who found reductions in Covid-19 associated with HCQ when all patients (not stratified by time) below 50 years were considered (RR 0.71; 95%CI 0.48-1.05, p=0.089), with a significant reduction associated with HCQ in the cohort younger than 50 years experiencing a high-risk exposure (RR 0.63; 95%CI 0.41-0.95, p= 0.0293). HCQ-associated reductions in Hazard/Risk ratios of a similar order of magnitude can be seen when comparing similar age strata in the PEP (Early cohort) and PrEP studies (Table S 6).

We verified the finding of Watanabe [28] who, not having the raw data, closely estimated from Figure 2 of the PEP study the percentage of patients becoming symptomatic after the full five days of treatment. Table S 7 shows that after removing subjects attaining a Covid-19 positive outcome by Day 5, there was a 41% reduction in Covid-19 associated with HCQ (RR 0.59, 95%CI 0.32-1.07, p=0.085). When stratified by intervention lag there were HCQ-associated reductions (not statistically significant) in the Early cohort (RR 0.42, 95%CI 0.18-0.99, p=0.056, NNT=20.2), and to a lesser extent in the Late cohort (RR 0.86, 95%CI 0.36-2.07, p=0.82).

*Severity of symptoms reported on day 14 from drug receipt*
We did not detect differences in the severity (median, IQR, n) of symptoms associated with HCQ reported on day 14 from drug receipt for both Early (HCQ 3.13; 1.95-4.58, n=10 vs. placebo 3.0; 1.9-4.1, n=21) and Late prophylaxis (HCQ 2.5; 1.65-4.1, n=23 vs. placebo 2.4; 1.4-5.1, n=13). This result is limited by the small number of subjects (n=67) reporting symptom scores.

*Provision of outcome data, adherence to study drug and use of folate*
Considering only the "Responding Population" (Table S 8), the effect associated with HCQ on the development of Covid-19 was similar to that seen in the whole population (Table S 9). Any small effect associated with HCQ observed (RR 0.82) was attenuated when only those subjects who were fully (RR 0.93) and fully or partially (RR 0.89) adherent to study medication were considered (Table S 9).

We examined how using folate as placebo might influence outcomes. The "no folate control" cohort had a slightly reduced (RR 0.93) development of Covid-19 compared with the "folate only placebo" cohort (Table S 10), resulting in a small change in the estimation of the effect associated with HCQ in the overall Responding Population. Stratifying these data by time reveals no discernible effect of the folate placebo (Table S 11). Combining the "folate only placebo" and "no folate control" cohorts, there was a directionally similar effect associated with HCQ (full plus partial adherence) to that observed in the ITT population for the Early prophylaxis cohort (RR 0.65, 95%CI 0.39-1.08; p=0.11).

*Ex-protocol use of zinc and Vitamin C*

The doses, duration of use, or reasons for self-medication with zinc or Vitamin C are unknown. The number of subjects was small in most of the sub-groups representing the different combinations of use of both agents, the largest of which (n=504) reported taking neither agent. Time stratified data for these subjects reveal an effect similar to that found in the ITT population, but without achieving statistical significance (Table S 12).

*Influence of co-morbidity on HCQ-associated outcomes*

Small population sizes (Table S 13) within the subgroups representing the three most frequent co-morbidities (hypertension, asthma, diabetes) prompt cautious analysis. However, for subjects reporting no co-existing conditions (72.6% of population) there was a reduction signal in Covid-19 associated with HCQ in the whole cohort (RR 0.7, 95%CI 0.46-1.06, p=0.094). Confirming the analysis by Luco, [27] when asthma and "other" co-morbidities are excluded, there was a similar signal (RR 0.72, 95%CI 0.49-1.05, p=0.097). Removing only asthmatic subjects yields a slightly weaker signal (RR 0.77, 95%CI 0.53-1.11, p=0.198). It must be noted that the incidence of Covid-19 observed for the asthma and the "other" co-morbidity sub-groups (6.5%) was much lower than that for the other groups (10.4-15.9%).

Stratifying by time and with no effects associated with Late HCQ prophylaxis, these trends achieved statistical significance associated with Early HCQ prophylaxis with no co-morbidity (RR 0.49, 95%CI 0.27-0.88, p=0.015, NNT 10.2), excluding asthma and "other" co-morbidity (RR 0.52, 95%CI 0.3-0.9, p=0.023) or excluding just subjects with asthma (RR 0.54, 95%CI 0.32-0.92, p=0.026).

Stratifying by age and considering only subjects with no co-morbidities, reveals a stronger response associated with Early HCQ prophylaxis for the younger (18-45) age group (RR 0.44, 95%CI 0.23-0.85, p=0.016). Although a stronger signal was observed in older subjects (>45 years) (RR 0.7, 95%CI 0.21-2.3, p= 0.739) that had not been seen in other age-related stratifications, this did not reach statistical significance.

*Sensitivity analysis*

Subsequent to the pre-printed version (December 12, 2020) of our manuscript, the original authors, in responding to questions regarding a companion study, provided two novel analyses [26] from their PEP study involving the "Days from High Risk Exposure to Enrollment" and "Days from Last Exposure to Study Drug Start." Although both of these analyses are flawed, they provide the basis for two sensitivity analyses.

*Sensitivity analysis #1. Mitigating effect of possibly higher placebo rate on individual days*

We note in the main body of this manuscript, an event rate for the placebo arm at numbered day 4 (18.9%) that is apparently higher than for other days. Although matched by a commensurate increase in the event rate for the HCQ arm, in an attempt to mitigate the effect of what might be an aberrant value, we mirrored the approach of the original authors to use a pooled placebo cohort in a sensitivity analysis.

In the first of their secondary analyses (Figure 1A of [26]), the original authors compared the Covid-19 event rate for "Days from High Risk Exposure to Enrollment" with the pooled placebo cohort. This is valueless and flawed in two respects: (1) it uses "Days from" instead of "Day on" nomenclature; (2) it does not consider actual shipping times, a problem that was known at least 2 months prior to the publication of this secondary analysis on December 4 2020.

Although not shown in this analysis, the daily placebo event rates are 12.7%, 14.5%, and 12.4% respectively for numbered days 1,3 and 4, with a higher value (17%) at day 2. Although pooling of placebo data across time periods obscures information about the trajectory of Covid-19 illness after exposure, this approach has some value in a sensitivity analysis to mitigate the effects of possibly aberrant individual values for absolute risk.

With these limitations, we applied this approach of using a pooled placebo cohort to compare the event rates for individual days using the intervention lag from highest risk exposure to drug receipt. This analysis (Table S 14, Figure S 1) shows the same trends as we have described in our main analysis (Table 1), with reductions seen for early but not Late cohorts. Using 1-3 days elapsed time of intervention lag (numbered days 2-4) for the Early prophylaxis cohort, there is a 32.5% reduction in Covid-19 associated with HCQ (RR 0.674; 95%CI 0.42-1.1, p=0.124). Taking only 1-2 days elapsed time intervention lag, we obtain a 42.9% reduction (RR 0.571, 95%CI 0.30-

1.08, p=0.09). TTable S *14*his analysis appears to reveal a strong regression line (p=0.033) of Covid-19 reduction (as 1-RR) and intervention lag (Figure S 1). Thus, even using the approach (using a pooled placebo cohort) advocated by the original authors to mitigate the effects of possibly outlying values in the placebo event rate, still yields support for our overall conclusions suggesting that early intervention with HCQ is an effective strategy.

*Sensitivity analysis #2. Time from Last Exposure*

In the second of their secondary analyses (Figure 1B of [26]), the original authors also used a pooled placebo cohort to examine Covid-19 event rate for “Days from Last Exposure to Study Drug Start”

This variable was not described in the original publication or protocol and was not provided in the released datasets. We suggest, as in a similar PEP study, [32] that the lag from last exposure provides an extremely limited basis for interpretation, as potentially pathogenic exposure has clearly occurred before the “last” exposure. A further problem is that it is unclear whether the “Days from” or “Day on” nomenclature is being used. Lastly, rather than report the data for the same ITT cohort used in the original primary analysis [33], a modified intent-to-treat analysis (mITT) was reported *“excluding those who never started the study medicine or withdrew from the trial.”*

With these caveats, and without publicly available patient-level data for this parameter, we extracted sub-group values from the secondary publication [26] as described in Table S 15. Pooling data for days 1-3 and comparing with the pooled (all times) placebo values, we find a reduction in Covid-19 associated with HCQ when given on days 1-3 (unknown nomenclature) after last exposure from 15.2% to 11.2% (RR 0.74, 95%CI 0.48-1.14, p=0.179). Although not directly comparable, the mITT population used by the authors appears somewhat similar to the population we used in our “Responding Population” which yielded results of a similar order of magnitude (Table S *11*).

Table S 1: Demographic and clinical characteristics, stratified into Early and Late Cohorts

The data for the original cohort recreates data from the original paper, for comparison and quality control purposes. Several variables have been added. The data are stratified into the Early (1-3 days) and Late (4-6 days) post exposure prophylaxis cohorts. See note in Table 1 regarding the nomenclature for "days." (I/S/%) - Shown in parentheses are interquartile ranges (1st and 3rd quartile), or standard deviations where indicated. All other parentheses indicate the percent contribution to the cohort total.

| | **Original Cohort** | | | | **Early (<= 3 days from exposure)** | | | | **Late Cohort (4-6 days from exposure)** | | | |
|---|---|---|---|---|---|---|---|---|---|---|---|---|
| **Characteristic** | **HCQ** | | **Placebo** | | **HCQ** | | **Placebo** | | **HCQ** | | **Placebo** | |
| **n** | **414** | | **407** | | **208** | | **218** | | **205** | | **189** | |
| **Age** | | **(I/S/%))** | | **(I/S/%))** | | **(I/S/%))** | | **(I/S/%))** | | **(I/S/%))** | | **(I/S/%))** |
| Median Age (IQR) | 41 | (33-51) | 40 | (32-50) | 40 | (33-49) | 39 | (32-49) | 42 | (32-52) | 41 | (33-51) |
| Average age (SD) | 42.3 | (12.7) | 41.8 | (12.0) | 42.1 | (12.3) | 41.1 | (11.7) | 42.6 | (13.2) | 42.7 | (12.3) |
| **Age distribution (%) of cohort)** | | | | | | | | | | | | |
| Age 18-35 | 151 | (36%) | 145 | (36%) | 75 | (36%) | 84 | (39%) | 76 | (37%) | 61 | (32%) |
| Age 35-50 | 159 | (38%) | 171 | (42%) | 89 | (43%) | 93 | (43%) | 69 | (34%) | 78 | (41%) |
| Age >50 | 104 | (25%) | 91 | (22%) | 44 | (21%) | 41 | (19%) | 60 | (29%) | 50 | (26%) |
| Median weight (kg) | 75 | (64-86) | 76 | (64-91) | 75 | (64-89) | 77 | (64-93) | 75 | (63-84) | 74 | (62-86) |
| **Biologic Sex** | | | | | | | | | | | | |
| Female | 218 | (52.7%) | 206 | (50.6%) | 102 | (49.0%) | 106 | (48.6%) | 116 | (56.6%) | 100 | (52.9%) |
| Male | 192 | (46.4%) | 197 | (48.4%) | 104 | (50.0%) | 111 | (50.9%) | 87 | (42.4%) | 86 | (45.5%) |
| Not stated | 4 | (1.0%) | 4 | (1.0%) | 2 | (1.0%) | 1 | (0.5%) | 2 | (1.0%) | 3 | (1.6%) |
| **Ethnicity (all that apply)** | | | | | | | | | | | | |
| White or Caucasian | 245 | (59.2%) | 262 | (64.4%) | 128 | (61.5%) | 146 | (67.0%) | 117 | (57.1%) | 116 | (61.4%) |
| Black or African American | 19 | (4.6%) | 18 | (4.4%) | 9 | (4.3%) | 11 | (5.0%) | 10 | (4.9%) | 7 | (3.7%) |
| Asian | 92 | (22.2%) | 83 | (20.4%) | 45 | (21.6%) | 37 | (17.0%) | 47 | (22.9%) | 46 | (24.3%) |
| Native Hawaiian or Pacific Islander | 2 | (0.5%) | 2 | (0.5%) | 2 | (1.0%) | 1 | (0.5%) | 0 | (0.0%) | 1 | (0.5%) |
| Hispanic or Latino | 22 | (5.3%) | 23 | (5.7%) | 10 | (4.8%) | 11 | (5.0%) | 12 | (5.9%) | 12 | (6.3%) |
| Native American or Alaska Native | 2 | (0.5%) | 1 | (0.2%) | 1 | (0.5%) | 0 | (0.0%) | 0 | (0.0%) | 1 | (0.5%) |
| Middle Eastern | 11 | (2.7%) | 2 | (0.5%) | 4 | (1.9%) | 1 | (0.5%) | 7 | (3.4%) | 1 | (0.5%) |
| South Asian | 18 | (4.3%) | 20 | (4.9%) | 9 | (4.3%) | 12 | (5.5%) | 9 | (4.4%) | 8 | (4.2%) |
| Other | 6 | (1.4%) | 3 | (0.7%) | 2 | (1.0%) | 3 | (1.4%) | 4 | (2.0%) | 0 | (0.0%) |
| **Current Smoker** | | | | | | | | | | | | |
| Current Smoker | 15 | (3.6%) | 12 | (2.9%) | 6 | (2.9%) | 9 | (4.1%) | 8 | (3.9%) | 3 | (1.6%) |
| Non-smoker | 395 | (95.4%) | 391 | (96.1%) | 200 | (96.2%) | 208 | (95.4%) | 195 | (95.1%) | 183 | (96.8%) |
| Not stated | 4 | (1.0%) | 4 | (1.0%) | 2 | (1.0%) | 1 | (0.5%) | 2 | (1.0%) | 3 | (1.6%) |

| | | | | | | | | | | | | |
|---|---|---|---|---|---|---|---|---|---|---|---|---|
| **Country** | | | | | | | | | | | | |
| Canada | 10 | (2.4%) | 11 | (2.7%) | 7 | (3.4%) | 10 | (4.6%) | 3 | (1.5%) | 1 | (0.5%) |
| United States | 404 | (97.6%) | 396 | (97.3%) | 201 | (96.6%) | 208 | (95.4%) | 202 | (98.5%) | 188 | (99.5%) |
| **Regularly Taking Any of These Medications** | | | | | | | | | | | | |
| Losartan | 14 | (3.4%) | 15 | (3.7%) | 8 | (3.8%) | 9 | (4.1%) | 6 | (2.9%) | 6 | (3.2%) |
| Aspirin | 10 | (2.4%) | 13 | (3.2%) | 4 | (1.9%) | 10 | (4.6%) | 6 | (2.9%) | 3 | (1.6%) |
| Ibuprofen/naproxen | 8 | (1.9%) | 8 | (2.0%) | 1 | (0.5%) | 4 | (1.8%) | 7 | (3.4%) | 4 | (2.1%) |
| Tylenol | 8 | (1.9%) | 11 | (2.7%) | 2 | (1.0%) | 5 | (2.3%) | 6 | (2.9%) | 6 | (3.2%) |
| No medications | 290 | (70.0%) | 269 | (66.1%) | 133 | (63.9%) | 125 | (57.3%) | 156 | (76.1%) | 144 | (76.2%) |
| Taking zinc in study | 100 | (24.2%) | 85 | (20.9%) | 47 | (22.6%) | 46 | (21.1%) | 53 | (25.9%) | 39 | (20.6%) |
| Taking Vitamin C in study | 140 | (33.8%) | 130 | (31.9%) | 68 | (32.7%) | 71 | (32.6%) | 71 | (34.6%) | 59 | (31.2%) |
| **Contact type** | | | | | | | | | | | | |
| HCW | 275 | (66.4%) | 270 | (66.3%) | 147 | (70.7%) | 152 | (69.7%) | 128 | (62.4%) | 118 | (62.4%) |
| Household | 125 | (30.2%) | 120 | (29.5%) | 54 | (26.0%) | 56 | (25.7%) | 71 | (34.6%) | 64 | (33.9%) |
| High-risk exposure | 365 | (88.2%) | 354 | (87.0%) | 186 | (89.4%) | 180 | (82.6%) | 179 | (87.3%) | 174 | (92.1%) |
| No PPE worn | 258 | (62.3%) | 237 | (58.2%) | 126 | (60.6%) | 116 | (53.2%) | 132 | (64.4%) | 121 | (64.0%) |
| **Numbered Day of receipt of study drug (Day 1 = exposure) as % of cohort.** | | | | | | | | | | | | |
| 1 | 0 | | 0 | | 0 | | 0 | | | | | |
| 2 | 32 | (7.7%) | 20 | (4.9%) | 32 | (15.4%) | 20 | (9.2%) | | | | |
| 3 | 91 | (22.0%) | 92 | (22.6%) | 91 | (43.8%) | 92 | (42.2%) | | | | |
| 4 | 85 | (20.6%) | 106 | (26.0%) | 85 | (40.9%) | 106 | (48.6%) | | | | |
| 5 | 123 | (29.8%) | 119 | (29.2%) | | | | | 123 | (60.0%) | 119 | (63.0%) |
| 6 | 62 | (15.0%) | 62 | (15.2%) | | | | | 62 | (30.2%) | 62 | (32.8%) |
| 7 | 20 | (4.8%) | 8 | (2.0%) | | | | | 20 | (9.8%) | 8 | (4.2%) |
| **Coexisting conditions/ Chronic health conditions (all that apply)** | | | | | | | | | | | | |
| None | 306 | (73.9%) | 290 | (71.3%) | 159 | (76.4%) | 156 | (71.6%) | 146 | (71.2%) | 134 | (70.9%) |
| High blood pressure | 51 | (12.3%) | 48 | (11.8%) | 25 | (12.0%) | 28 | (12.8%) | 26 | (12.7%) | 20 | (10.6%) |
| Asthma | 31 | (7.5%) | 31 | (7.6%) | 15 | (7.2%) | 15 | (6.9%) | 16 | (7.8%) | 16 | (8.5%) |
| Diabetes | 12 | (2.9%) | 16 | (3.9%) | 6 | (2.9%) | 7 | (3.2%) | 6 | (2.9%) | 9 | (4.8%) |
| Cardiovascular disease | 4 | (1.0%) | 2 | (0.5%) | 4 | (1.9%) | 1 | (0.5%) | 0 | (0.0%) | 1 | (0.5%) |
| Cancer or malignancy | 1 | (0.2%) | 2 | (0.5%) | 0 | (0.0%) | 2 | (0.9%) | 1 | (0.5%) | 0 | (0.0%) |
| Chronic kidney disease | 0 | (0.0%) | 3 | (0.7%) | 0 | (0.0%) | 3 | (1.4%) | 0 | (0.0%) | 0 | (0.0%) |
| Other chronic lung disease | 3 | (0.7%) | 0 | (0.0%) | 1 | (0.5%) | 0 | (0.0%) | 2 | (1.0%) | 0 | (0.0%) |
| Chronic liver disease | 0 | (0.0%) | 0 | (0.0%) | 0 | (0.0%) | 0 | (0.0%) | 0 | (0.0%) | 0 | (0.0%) |
| HIV | 1 | (0.2%) | 0 | (0.0%) | 1 | (0.5%) | 0 | (0.0%) | 0 | (0.0%) | 0 | (0.0%) |
| Transplant recipient | 0 | (0.0%) | 1 | (0.2%) | 0 | (0.0%) | 1 | (0.5%) | 0 | (0.0%) | 0 | (0.0%) |

| | | | | | | | | | | | | |
|---|---|---|---|---|---|---|---|---|---|---|---|---|
| Corticosteroids, chemotherapy, immunosuppressive | 2 | (0.5%) | 1 | (0.2%) | 0 | (0.0%) | 1 | (0.5%) | 2 | (1.0%) | 0 | (0.0%) |
| Hepatitis B or C | 1 | (0.2%) | 0 | (0.0%) | 0 | (0.0%) | 0 | (0.0%) | 1 | (0.5%) | 0 | (0.0%) |
| Other | 25 | (6.0%) | 31 | (7.6%) | 13 | (6.3%) | 16 | (7.3%) | 12 | (5.9%) | 15 | (7.9%) |

Table S 2: Clarification of Exposure Risk Definition

The PEP [33] publication defines the risk of exposure as household or occupational exposure to someone with confirmed Covid-19 at a distance of less than 6 ft for more than 10 minutes as:

- High-risk exposure: wearing neither a face mask nor an eye shield
- Moderate-risk exposure: wearing a face mask but no eye shield

The principal investigator (personal communication) informed us that due to changing CDC HCW risk guidance, the risk score changed over time (March 17, March 19, and April 3, 2020) and that there was more nuance than was captured in the database. There was also discussion with some of the participants regarding their risk definition (example – HCW who wore PPE but then removed it in the presence of the patient). From March 19 onward, the risk definition was:

| **Contact Type** | **Risk type** | **Distance/Time** | **PPE** | **Eye protection** | **Face protection** |
|---|---|---|---|---|---|
| Household | High | < 6ft + > 10 min | None | | |
| Household | Moderate | < 6ft + > 10 min | Any | | |
| Household | Low | > 6ft or < 10 min | | | |
| HCW | High | < 6ft + > 10 min | "less than optimal" | | |
| HCW | High | < 6ft + > 10 min | | No | No |
| HCW | High | < 6ft + > 10 min | | No | Yes |
| HCW | High | < 6ft + > 10 min | | Yes | No |
| HCW | Moderate | < 6ft + > 10 min | Full | | |
| HCW | Moderate | < 6ft + > 10 min | | Yes | Yes |
| HCW | Low | > 6ft or < 10 min | | | |

Table S 3: Distribution of days of drug receipt within strata for study day of enrollment

| | **Numbered Day of drug receipt** | | | | | | | |
|---|---|---|---|---|---|---|---|---|
| **Numbered Day of Enrollment** | **1** | **2** | **3** | **4** | **5** | **6** | **7** | Total |
| **1** | 0 | 52 | 70 | 18 | 0 | 0 | 0 | 140 |
| **2** | 0 | 0 | 113 | 67 | 26 | 0 | 0 | 206 |
| **3** | 0 | 0 | 0 | 98 | 94 | 23 | 0 | 215 |
| **4** | 0 | 0 | 0 | 8 | 122 | 101 | 28 | 259 |

For each of the enrollment days described in the original report, the numbered days on which drug receipt occurred are shown. Day 1 = day of exposure. See note in Table 1 regarding the nomenclature for "days." Note time-related data for one subject (#308) are missing. In the original, the 10/6 and the 10/30 datasets, data for subject #308 for the variables describing the time from exposure to enrollment or drug receipt were missing. Although data for time between enrollment and shipping (1.58 days) were provided in the 9/9 dataset for this subject, it was not possible to assign this subject to any time stratum. We have therefore retained the exclusion of this subject from the applicable analyses, as in the original work.

Table S 4: Stratification of effect associated with hydroxychloroquine by gender based on time from exposure to drug receipt (ITT population)

| | Hydroxychloroquine | | | Placebo | | | | | |
|---|---|---|---|---|---|---|---|---|---|
| | **n Pos** | **N Total** | **%Pos** | **n Pos** | **N Total** | **%Pos** | **RR** | **CI Low** | **CI Up** |
| **Early prophylaxis 1-3 days post exposure (numbered days 2-4)** | | | | | | | | | |
| **Male** | 7 | 104 | 6.7% | 14 | 111 | 12.6% | 0.53 | 0.22 | 1.27 |
| **Female** | 13 | 102 | 12.7% | 21 | 106 | 19.8% | 0.64 | 0.34 | 1.22 |
| **Late prophylaxis 4-6 days post-exposure (numbered days 5-7)** | | | | | | | | | |
| **Male** | 12 | 87 | 13.8 | 10 | 86 | 11.6% | 1.19 | 0.54 | 2.60 |
| **Female** | 17 | 116 | 14.7% | 12 | 100 | 12.0% | 1.22 | 0.61 | 2.43 |

The number (and percent) of subjects with a Covid-19 positive outcome are shown for each group along with the total number of subjects for that group, stratified by time from exposure to drug receipt. The elapsed time range in days is shown for Early and Late cohorts. See note in Table 1 regarding the nomenclature for “days.” See note in Table S 3 regarding tallies for time-stratified data.

Table S 5: Stratification of effect associated with hydroxychloroquine by exposure risk level based on time from exposure to drug receipt (ITT population) and contact type

A: Exposure risk level and time to drug receipt

| | Hydroxychloroquine | | | Plac | | | | | | | |
|---|---|---|---|---|---|---|---|---|---|---|---|
| | nPos | N Total | %Pos | nPos | N Total | %Pos | RR | CI Low | CI Up | NNT | p |
| **All times** | | | | | | | | | | | |
| High | 43 | 365 | 11.8% | 54 | 354 | 15.3% | 0.77 | 0.53 | 1.12 | 28.8 | 0.191 |
| Moderate | 6 | 48 | 12.5% | 4 | 53 | 7.5% | 1.66 | 0.50 | 5.52 | | |
| **Early prophylaxis 1-3 days post exposure (numbered days 2-4)** | | | | | | | | | | | |
| High | 16 | 186 | 8.6% | 32 | 180 | 17.8% | 0.48 | 0.28 | 0.85 | 10.9 | 0.013 |
| Moderate | 4 | 22 | 18.2% | 4 | 38 | 10.5% | 1.73 | 0.48 | 6.23 | | |
| **Late prophylaxis 4-6 days post-exposure (numbered days 5-7)** | | | | | | | | | | | |
| High | 27 | 179 | 15.1% | 22 | 174 | 12.6% | 1.19 | 0.71 | 2.01 | | |
| Moderate | 2 | 26 | 7.7% | 0 | 15 | 0.0% | | | | | |

**B: Contact type and e**xposure risk level

| | Hydroxychloroquine | | | Placebo | | | | | | |
|---|---|---|---|---|---|---|---|---|---|---|
| | n Pos | N Total | %Pos | n Pos | N Total | %Pos | RR | CI Low | CI Up | p |
| **Household: % Mod** | | | 6.4% | | | 5.8% | | | | |
| **High** | 16 | 117 | 13.7% | 25 | 113 | 22.1% | 0.62 | 0.35 | 1.09 | 0.12 |
| **Moderate** | 2 | 8 | 25.0% | 0 | 7 | 0.0% | | | | |
| **Health Care Worker: % Mod** | | | 13.1% | | | 15.2% | | | | |
| **High** | 27 | 239 | 11.3% | 29 | 229 | 12.7% | 0.89 | 0.55 | 1.46 | 0.67 |
| **Moderate** | 4 | 36 | 11.1% | 4 | 41 | 9.8% | 1.14 | 0.31 | 4.23 | |

Shown as %Mod is the percent contribution of moderate risk cases to the total number of cases, by treatment and exposure type. The risk level is unknown for subject #308.

The number (and percent) of subjects with a Covid-19 positive outcome are shown for each group along with the total number of subjects for that group, stratified by time from exposure to drug receipt. The elapsed time range in days is shown for Early and Late cohorts. See note in Table 1 regarding the nomenclature for “days.” See note in Table S 3 regarding tallies for time-stratified data.

Table S 6: Comparison of HCQ-associated effects by age in PEP and PrEP studies

| **Age Range** | | **Hydroxychloroquine** | | | **Placebo** | | | | **95% CI** | | | |
|---|---|---|---|---|---|---|---|---|---|---|---|---|
| **From** | **To** | **n Pos** | **N Total** | **%Pos** | **n Pos** | **N Total** | **%Pos** | **HR/ RR** | **Lower** | **Upper** | **NNT** | **p** |
| **Early PEP - prophylaxis 1-3 days post exposure (numbered days 2-4)** | | | | | | | | | | | | |
| **>18** | **<=45** | 14 | 140 | 10.0% | 28 | 150 | 18.7% | 0.54 | 0.29 | 0.97 | 11.5 | 0.045 |
| **>18** | **<=50** | 17 | 164 | 10.4% | 35 | 177 | 19.8% | 0.52 | 0.31 | 0.90 | 10.6 | 0.016 |
| **PrEP Study – Pre-exposure Prophylaxis** | | | | | | | | | | | | |
| **>18** | **<=39** | 23 | 420 | 5.5% | 23 | 232 | 9.9% | 0.55 | 0.32 | 0.96 | 22.5 | 0.038 |
| **>18** | **<=50** | 43 | 769 | 5.6% | 34 | 404 | 8.4% | 0.66 | 0.43 | 1.02 | 35.4 | 0.082 |

The number (and percent) of subjects with a Covid-19 positive outcome are shown for each group along with the total number of subjects for that group. The data for the PrEP study are from the pre-printed [29] and published [30] versions for the two dose groups combined. HR, RR and 95%CI values are calculated without adjustment. See note in Table 1 regarding the nomenclature for “days.”

Table S 7: Analysis considering only patients becoming positive after five days of treatment

| Numbered | Hydroxychloroquine | | | Placebo | | | | 95% CI | | | |
|---|---|---|---|---|---|---|---|---|---|---|---|
| **Day** | **nPos** | **N Total** | **%Pos** | **n Pos** | **N Total** | **%Pos** | **RR** | **Lower** | **Upper** | **NNT** | **P** |
| **1** | 0 | 0 | | 0 | 0 | | | | | | |
| **2** | 0 | 30 | 0.0% | 2 | 20 | 10.0% | 0 | | | | |
| **3** | 4 | 87 | 4.6% | 5 | 83 | 6.0% | 0.76 | 0.21 | 2.74 | | |
| **4** | 3 | 78 | 3.8% | 10 | 96 | 10.4% | 0.37 | 0.11 | 1.30 | | |
| **5** | 7 | 113 | 6.2% | 7 | 112 | 6.3% | 0.99 | 0.36 | 2.73 | | |
| **6** | 2 | 57 | 3.5% | 3 | 57 | 5.3% | 0.67 | 0.12 | 3.84 | | |
| **7** | 0 | 15 | 0.0% | 0 | 8 | 0.0% | | | | | |
| **Early prophylaxis 1-3 days post exposure (numbered days 2-4)** | | | | | | | | | | | |
| | 7 | 195 | 3.6% | 17 | 199 | 8.5% | 0.42 | 0.18 | 0.99 | 20.2 | 0.056 |
| **Late prophylaxis 4-6 days post-exposure (numbered days 5-7)** | | | | | | | | | | | |
| | 9 | 185 | 4.9% | 10 | 177 | 5.6% | 0.86 | 0.36 | 2.07 | 127.4 | 0.82 |
| **Prophylaxis all times 1-6 days post-exposure (numbered days 2-7)** | | | | | | | | | | | |
| **All** | 16 | 380 | 4.2% | 27 | 376 | 7.2% | 0.59 | 0.32 | 1.07 | 33.7 | 0.085 |

The number (and percent) of subjects with a Covid-19 positive outcome are shown for each group along with the total number of subjects for that group, stratified by time from exposure to drug receipt. The Days shown are the numbered days on which study drug was received, with Day 1 = day of reported high risk exposure. See note in Table 1 regarding the nomenclature for “days.”

Table S 8: Summary of subjects forming “Responding Population”

| | Total | HCQ | Placebo |
|---|---|---|---|
| Original ITT Cohort | 821 | 414 | 407 |
| *Excluded will be subjects:* | | | |
| Withdrew Consent | 8 | 4 | 4 |
| LTF, no survey data per Table S1 in PEP study | 55 | 25 | 30 |
| LTF, noted as "Some Survey Data", but no symptoms | 9 | 5 | 4 |
| **Totals for exclusion** | **72** | **34** | **38** |
| **Total included in Responding Population** | 749 | 380 | 369 |

Of the 88 LTF patients, 52 were reported as not completing any surveys and were unresponsive to follow up. Another 36 had: some survey data with vital status after day 14 known (16), no survey with vital status after day 14 known (3) or no survey with vital status after day 14 unknown (17). We examined the 33 patients noted as having some survey data and found that there were 9 with no symptom data. There was a total of 72 patients with no symptom data at all which we excluded from the Responding Population. The remaining 24 patients had incomplete symptom data for days 3, 5, 10 and 14 in various combinations and we considered their outcomes as includable using the “last observation carried forward” (LOCF) method using the endpoint adjudication of the original authors.

Table S 9: Effect of adherence to study drug on development of Covid-19 within Responding Population (RP)

| | **Hydroxychloroquine** | | | **Placebo** | | | | | |
|---|---|---|---|---|---|---|---|---|---|
| **Adherence** | **n Pos** | **N Total** | **%** | **n Pos** | **N Total** | **%** | **RR** | **CI Low** | **CI Up** |
| ITT Cohort, as published | 49 | 414 | 11.8% | 58 | 407 | 14.3% | 0.83 | 0.58 | 1.18 |
| All subjects (RP) | 49 | 380 | 12.9% | 58 | 369 | 15.7% | 0.82 | 0.58 | 1.17 |
| Fully adherent (RP) | 43 | 312 | 13.8% | 50 | 336 | 14.9% | 0.93 | 0.64 | 1.35 |
| Partially adherent (RP) | 4 | 36 | 11.1% | 3 | 12 | 25.0% | 0.44 | 0.12 | 1.71 |
| Not adherent (RP) | 2 | 32 | 6.3% | 5 | 21 | 23.8% | | | |
| Fully + Partial adherence (RP) | 47 | 348 | 13.5% | 53 | 348 | 15.2% | 0.89 | 0.62 | 1.28 |

The number (and percent) of subjects with a Covid-19 positive outcome are shown for each group along with the total number of subjects for that group, stratified by adherence to study medication. The data from the ITT cohort from the original paper are shown for reference.

Table S 10: Effect associated with folate placebo in Responding Population

| | **Hydroxychloroquine** | | | **Placebo** | | | | | |
|---|---|---|---|---|---|---|---|---|---|
| | **n Pos** | **N Total** | **%** | **n Pos** | **N Total** | **%** | **RR** | **CI Low** | **CI Up** |
| HCQ vs. Folate only placebo | 47 | 348 | 13.5% | 51 | 337 | 15.1% | 0.89 | 0.62 | 1.29 |
| HCQ vs. no Folate control | 47 | 348 | 13.5% | 9 | 64 | 14.1% | 0.96 | 0.50 | 1.86 |
| HCQ vs. Combined control | 47 | 348 | 13.5% | 60 | 401 | 15.0% | 0.90 | 0.63 | 1.29 |
| | **No folate** | | | **Folate Placebo** | | | | | |
| No Folate control vs. Folate only Placebo | 9 | 64 | 14.1% | 51 | 337 | 15.1% | 0.93 | 0.48 | 1.79 |

Shown is the percent (n/N) of subjects with Covid-19 positive outcome for the fully and partially adherent subgroups combined.

Table S 11: Effect associated with folate placebo in Responding Population (fully and partially adherent study subjects), stratified by time and gender.

| | n Pos | N Total | % Pos | RR | 95% CI Lower | Upper | p |
|---|---|---|---|---|---|---|---|
| **Early prophylaxis 1-3 days post exposure (numbered days 2-4)** | | | | | | | |
| **Male + Female** | | | | | | | |
| HCQ | 19 | 173 | 11.0% | | | | |
| Folate Placebo | 31 | 183 | 16.9% | 0.65 | 0.38 | 1.10 | p=0.127 |
| No Folate | 6 | 35 | 17.1% | 0.64 | 0.28 | 1.49 | |
| Combined Control | 37 | 218 | 17.0% | 0.65 | 0.39 | 1.08 | p=0.11 |
| **MALE** | | | | | | | |
| HCQ | 83 | 7 | 8.4% | | | | |
| Folate Placebo | 90 | 12 | 13.3% | 0.63 | 0.26 | 1.53 | |
| No Folate | 21 | 2 | 9.5% | 0.89 | 0.20 | 3.96 | |
| Combined Control | 111 | 14 | 12.6% | 0.67 | 0.28 | 1.58 | |
| **FEMALE** | | | | | | | |
| HCQ | 89 | 12 | 13.5% | | | | |
| Folate Placebo | 92 | 18 | 19.6% | 0.69 | 0.35 | 1.35 | |
| No Folate | 14 | 4 | 28.6% | 0.47 | 0.18 | 1.26 | |
| Combined Control | 106 | 22 | 20.8% | 0.65 | 0.34 | 1.24 | |
| **Late prophylaxis 4-6 days post-exposure (numbered days 5-7)** | | | | | | | |
| **Male + Female** | | | | | | | |
| HCQ | 28 | 174 | 16.1% | | | | |
| Folate Placebo | 20 | 154 | 13.0% | 1.24 | 0.73 | 2.11 | |
| No Folate | 3 | 29 | 10.3% | 1.56 | 0.51 | 4.79 | |
| Combined Control | 23 | 183 | 12.6% | 1.28 | 0.77 | 2.13 | |
| **MALE** | | | | | | | |
| HCQ | 77 | 11 | 14.3% | | | | |
| Folate Placebo | 73 | 9 | 12.3% | 1.16 | 0.51 | 2.63 | |
| No Folate | 12 | 2 | 16.7% | 0.86 | 0.22 | 3.40 | |
| Combined Control | 85 | 11 | 12.9% | 1.10 | 0.51 | 2.40 | |
| **FEMALE** | | | | | | | |
| HCQ | 95 | 17 | 17.9% | | | | |
| Folate Placebo | 79 | 11 | 13.9% | 1.29 | 0.64 | 2.58 | |
| No Folate | 17 | 1 | 5.9% | 3.04 | 0.43 | 21.37 | |
| Combined Control | 96 | 12 | 12.5% | 1.43 | 0.72 | 2.83 | |

RR - Risk Ratios vs. HCQ shown
Shown is the percent (n/N) of subjects with Covid-19 positive outcome for the fully and partially adherent subgroups combined, stratified by control group type and gender. See note in Table 1 regarding the nomenclature for “days.” See note in Table S 3 regarding tallies for time-stratified data.

Table S 12: Effect of ex-protocol use of Zinc or Vitamin C on effect associated with hydroxychloroquine stratified by time from exposure to drug receipt

Part A: Numbers of subjects reporting at two subject surveys to be taking zinc or Vitamin C.

| | **Vitamin C** | | | | |
|---|---|---|---|---|---|
| **Zinc** | **None** | **d5 only** | **d14 only** | **both** | **any** |
| **None** | 504 * | 24 | 49 | 59 * | 132 * |
| **d5 only** | 6 | 8 | 0 | 6 | 14 |
| **d14 only** | 19 | 1 | 32 | 13 | 46 |
| **both** | 22 * | 4 | 4 | 70 * | 78 * |
| **any** | 47* | 13 | 36 | 89 * | 138* |

Subject surveys were carried out on days 5 and 14 numbering from day of receipt of study drug. The number of subjects reporting taking either agent at either or both days 5 and 14 are shown. Detail for use of either agent at these time points was provided at our request in the 9/9 revision. The combinations marked with an asterisk * are subject to further analysis below, representing use of neither agent, both agents at both times either agent at any or both times time. Note that the total number of subjects reporting use of zinc is 185, which corrects a typographic error in the original paper.

Part B: Effect of ex-protocol use of zinc or vitamin C on effect associated with hydroxychloroquine stratified by time from exposure to drug receipt and by reported use of zinc or vitamin C.

The number (and percent) of subjects with a Covid-19 positive outcome are shown for each group along with the total number of subjects for that group, stratified by time from exposure to drug receipt (as Early to Late Prophylaxis cohorts).

| | | | **HCQ** | | | **Placebo** | | | | **95% CI** | |
|---|---|---|---|---|---|---|---|---|---|---|---|
| **Zinc** | **Vit C** | | **n Pos** | **N Total** | **%Pos** | **n Pos** | **N Total** | **%Pos** | **RR** | **CI Low** | **CI Up** |
| ITT population | | Early | 20 | 208 | 9.6% | 36 | 218 | 16.5% | 0.58 | 0.35 | 0.97 |
| ITT population | | Late | 29 | 205 | 14.1% | 22 | 189 | 11.6% | 1.22 | 0.72 | 2.04 |
| None | None | Early | 9 | 128 | 7.0% | 18 | 137 | 13.1% | 0.54 | 0.25 | 1.15 |
| None | None | Late | 16 | 120 | 13.3% | 10 | 119 | 8.4% | 1.59 | 0.75 | 3.35 |
| Both | Both | Early | 2 | 18 | 11.1% | 1 | 13 | 7.7% | 1.44 | 0.15 | 14.29 |
| Both | Both | Late | 3 | 19 | 15.8% | 2 | 20 | 10.0% | 1.58 | 0.30 | 8.43 |
| Both | None | Early | 1 | 6 | 16.7% | 1 | 3 | 33.3% | 0.50 | 0.05 | 5.51 |
| Both | None | Late | 1 | 5 | 20.0% | 1 | 8 | 12.5% | 1.60 | 0.13 | 20.22 |
| None | Both | Early | 2 | 11 | 18.2% | 3 | 13 | 23.1% | 0.79 | 0.16 | 3.90 |
| None | Both | Late | 0 | 15 | 0.0% | 4 | 19 | 21.1% | 0.00 | | |
| None | Any | Early | 4 | 33 | 12.1% | 10 | 35 | 28.6% | 0.42 | 0.15 | 1.22 |
| None | Any | Late | 5 | 32 | 15.6% | 7 | 31 | 22.6% | 0.69 | 0.25 | 1.95 |
| Any | None | Early | 1 | 12 | 8.3% | 2 | 10 | 20.0% | 0.42 | 0.04 | 3.95 |
| Any | None | Late | 3 | 14 | 21.4% | 1 | 11 | 9.1% | 2.36 | 0.28 | 19.66 |
| Both | Any | Early | 3 | 20 | 15.0% | 1 | 15 | 6.7% | 2.25 | 0.26 | 19.55 |
| Both | Any | Late | 3 | 23 | 13.0% | 2 | 20 | 10.0% | 1.30 | 0.24 | 7.04 |
| Any | Both | Early | 2 | 19 | 10.5% | 2 | 17 | 11.8% | 0.89 | 0.14 | 5.68 |
| Any | Both | Late | 4 | 27 | 14.8% | 4 | 26 | 15.4% | 0.96 | 0.27 | 3.45 |
| Any | Any | Early | 6 | 35 | 17.1% | 6 | 36 | 16.7% | 1.03 | 0.37 | 2.89 |
| Any | Any | Late | 5 | 39 | 12.8% | 4 | 28 | 14.3% | 0.90 | 0.26 | 3.05 |

Table S 13: Effect of comorbidities on effect associated with HCQ

Part A: Stratification by time only

| | nPos | N Total | % | nPos | N Total | % | RR | CI Low | CI Up | NNT | p |
|---|---|---|---|---|---|---|---|---|---|---|---|
| **Whole Cohort** | **Hydroxychloroquine** | | | **Placebo** | | | | | | | |
| All subjects | 49 | 414 | 11.8% | 58 | 407 | 14.3% | 0.83 | 0.58 | 1.18 | 41.4 | 0.351 |
| No co-morbidity | 34 | 306 | 11.1% | 46 | 290 | 15.9% | 0.70 | 0.46 | 1.06 | 21.0 | 0.094 |
| Hypertension | 7 | 51 | 13.7% | 5 | 48 | 10.4% | 1.32 | 0.45 | 3.87 | | |
| Asthma | 5 | 31 | 16.1% | 2 | 31 | 6.5% | 2.50 | 0.52 | 11.93 | | |
| Diabetes | 2 | 12 | 16.7% | 2 | 16 | 12.5% | 1.33 | 0.22 | 8.16 | | |
| Other | 4 | 25 | 16.0% | 2 | 31 | 6.5% | 2.48 | 0.49 | 12.45 | | |
| Excl. asthma or other | 40 | 361 | 11.1% | 54 | 349 | 15.5% | 0.72 | 0.49 | 1.05 | 22.8 | 0.097 |
| Exclude asthma only | 44 | 383 | 11.5% | 56 | 376 | 14.9% | 0.77 | 0.53 | 1.11 | 29.4 | 0.198 |
| **Early prophylaxis 1-3 days post exposure (numbered days 2-4)** | | | | | | | | | | | |
| All | 20 | 208 | 9.6% | 36 | 218 | 16.5% | 0.58 | 0.35 | 0.97 | 14.5 | 0.044 |
| No co-morbidity | 15 | 159 | 9.4% | 30 | 156 | 19.2% | 0.49 | 0.27 | 0.88 | 10.2 | 0.015 |
| Hypertension | 2 | 25 | 8.0% | 2 | 28 | 7.1% | 1.12 | 0.17 | 7.37 | | |
| Asthma | 2 | 15 | 13.3% | 1 | 15 | 6.7% | 2.00 | 0.20 | 19.78 | | |
| Diabetes | 1 | 6 | 16.7% | 0 | 7 | 0.0% | | | | | |
| Other | 1 | 13 | 7.7% | 1 | 16 | 6.3% | 1.23 | 0.08 | 17.83 | | |
| Excl. asthma or other | 17 | 181 | 9.4% | 34 | 189 | 18.0% | 0.52 | 0.30 | 0.90 | 11.6 | 0.023 |
| Exclude asthma only | 18 | 193 | 9.3% | 35 | 203 | 17.2% | 0.54 | 0.32 | 0.92 | 12.6 | 0.026 |
| **Late prophylaxis 4-6 days post exposure (numbered days 5-7)** | | | | | | | | | | | |
| All | 29 | 205 | 14.1% | 22 | 189 | 11.6% | 1.22 | 0.72 | 2.04 | | 0.548 |
| No co-morbidity | 19 | 147 | 12.9% | 16 | 134 | 11.9% | 1.08 | 0.58 | 2.02 | | 0.857 |
| Hypertension | 5 | 26 | 19.2% | 3 | 20 | 15.0% | 1.28 | 0.35 | 4.74 | | |
| Asthma | 3 | 16 | 18.8% | 1 | 16 | 6.3% | 3.00 | 0.35 | 25.87 | | |
| Diabetes | 1 | 6 | 16.7% | 2 | 9 | 22.2% | 0.75 | 0.09 | 6.55 | | |
| Other | 3 | 12 | 25.0% | 1 | 15 | 6.7% | 3.75 | 0.44 | 31.62 | | |
| Excl. asthma or other | 23 | 180 | 12.8% | 20 | 160 | 12.5% | 1.02 | 0.58 | 1.79 | | 1 |
| Exclude asthma only | 26 | 190 | 13.7% | 21 | 173 | 12.1% | 1.13 | 0.66 | 1.93 | | 0.755 |

Part B: Subjects reporting no comorbidities, stratified by age and time

| | nPos | N Total | % | nPos | N Total | % | RR | CI Low | CI Up | NNT | p |
|---|---|---|---|---|---|---|---|---|---|---|---|
| **Whole Cohort** | **Hydroxychloroquine** | | | **Placebo** | | | | | | | |
| All ages | 34 | 306 | 11.1% | 46 | 290 | 15.9% | 0.70 | 0.46 | 1.06 | 21.0 | 0.094 |
| 18-45 | 26 | 188 | 12.1% | 38 | 167 | 18.5% | 0.65 | 0.41 | 1.04 | 15.7 | 0.078 |
| 45-90 | 8 | 84 | 8.7% | 8 | 77 | 9.4% | 0.92 | 0.36 | 2.35 | 1 | 139.6 |
| **Early prophylaxis 1-3 days post exposure (numbered days 2-4)** | | | | | | | | | | | |
| All ages | 15 | 159 | 9.4% | 30 | 156 | 19.2% | 0.49 | 0.27 | 0.88 | 10.2 | 0.015 |
| 18-45 | 11 | 117 | 9.4% | 24 | 112 | 21.4% | 0.44 | 0.23 | 0.85 | 8.3 | 0.016 |
| 45-90 | 4 | 42 | 9.5% | 6 | 44 | 13.6% | 0.70 | 0.21 | 2.30 | 24.3 | 0.739 |
| **Late prophylaxis 4-6 days post exposure (numbered days 5-7)** | | | | | | | | | | | |
| All ages | 19 | 146 | 13.0% | 16 | 134 | 11.9% | 1.09 | 0.58 | 2.03 | | 0.857 |
| 18-45 | 15 | 96 | 15.6% | 14 | 93 | 15.1% | 1.04 | 0.53 | 2.03 | | 1 |
| 45-90 | 4 | 50 | 8.0% | 2 | 41 | 4.9% | 1.64 | 0.32 | 8.51 | | 0.686 |

Shown is the percent (n/N) of subjects with Covid-19 positive outcome for subjects reporting the co-existing condition specified, stratified by time from exposure to drug receipt. The elapsed time range in days is shown for Early and Late cohorts.

See note in Table 1 regarding the nomenclature for “days.” See note in Table S 3 regarding tallies for time-stratified data.

Table S 14: Sensitivity analysis, using pooled placebo cohort: *Stratification of effect associated with hydroxychloroquine based on time from exposure to drug receipt*

| **Numbered** | **Hydroxychloroquine** | | | **Placebo** | | | | **95% CI** | | |
|---|---|---|---|---|---|---|---|---|---|---|
| **Day** | **nPos** | **N Total** | **%Pos** | **n Pos** | **N Total** | **%Pos** | **RR** | **Lower** | **Upper** | |
| **1** | | | | | | | | | | |
| **2** | 2 | 32 | 6.3% | 58 | 407 | 14.3% | 0.439 | 0.112 | 1.714 | |
| **3** | 8 | 91 | 8.8% | 58 | 407 | 14.3% | 0.617 | 0.305 | 1.247 | |
| **4** | 10 | 85 | 11.8% | 58 | 407 | 14.3% | 0.826 | 0.440 | 1.549 | |
| **5** | 17 | 123 | 13.8% | 58 | 407 | 14.3% | 0.970 | 0.587 | 1.601 | |
| **6** | 7 | 62 | 11.3% | 58 | 407 | 14.3% | 0.792 | 0.379 | 1.656 | |
| **7** | 5 | 20 | 25.0% | 58 | 407 | 14.3% | 1.754 | 0.792 | 3.887 | |
| **Early prophylaxis 1-3 days post exposure (numbered days 2-4)** | | | | | | | | | | |
| | 20 | 208 | 9.6% | 58 | 407 | 14.3% | 0.675 | 0.418 | 1.09 | 0.124 |
| **Late prophylaxis 4-6 days post-exposure (numbered days 5-7)** | | | | | | | | | | |
| | 29 | 205 | 14.1% | 58 | 407 | 14.3% | 0.993 | 0.657 | 1.50 | |
| **Early prophylaxis 1-2 days post exposure (numbered days 2-3)** | | | | | | | | | | |
| | 10 | 123 | 8.1% | 58 | 407 | 14.3% | 0.571 | 0.301 | 1.082 | 0.09 |
| **Late prophylaxis 3-6 days post exposure (numbered days 4-7)** | | | | | | | | | | |
| | 39 | 290 | 13.4% | 58 | 407 | 14.3% | 0.944 | 0.647 | 1.376 | |

The number (and percent) of subjects with a Covid-19 positive outcome are shown for each group along with the total number of subjects for that group, stratified by time from exposure to drug receipt. The Days shown are the numbered days on which study drug was received, with Day 1 = day of reported high risk exposure. See note in Table 1 regarding the nomenclature for “days.” All comparisons are with the placebo cohort, pooled across all time periods. See also Figure S *1*.

Table S 15: Sensitivity analysis based "*Days from Last Exposure to Study Drug Start"* provided in Figure 1 of Nicol et al.

| Days | % Covid | n | nPos | nNeg | RR | 95%CI | p |
|---|---|---|---|---|---|---|---|
| 1 | 15.4 | 65 | 10.01 | 54.99 | | | |
| 2 | 9.7 | 113 | 10.96 | 102.04 | | | |
| 3 | 9.5 | 63 | 5.99 | 57.02 | | | |
| >=4 | 19.2 | 104 | 19.97 | 84.03 | | | |
| Days 1-3 combined | 11.2 | 241 | 26.96 | 214.04 | | | |
| Placebo | 15.2 | 349 | 53.05 | 295.95 | | | |
| Days 1-3 combined, rounded | 11.2 | 241 | 27 | 214 | 0.738 | 0.478-1.14 | 0.179 |
| Placebo, rounded | 15.2 | 349 | 53 | 296 | | | |

Data were extracted from in Figure 1 of Nicol et al. [26] which provides the number of subjects (n) in each time category and the percentage Covid-19 Compatible Illness (to one decimal place). The number of subjects with Covid-19 compatible illness was calculated (nPos) and rounded to the nearest integer. Due to ambiguity in the original figure, it is unclear whether the "Day on" or the "Days from" nomenclature is being used.

Data for Days 1-3 were combined, compared with the pooled placebo and an RR value computed. Note that values for the placebo arm for individual "day" stratum were not provided, nor are data for "Days from Last Exposure to Study Drug Start" provided in the released dataset.

Figure S 1: Sensitivity analysis, using pooled placebo cohort: Stratification of effect associated with hydroxychloroquine based on time from exposure to drug receipt

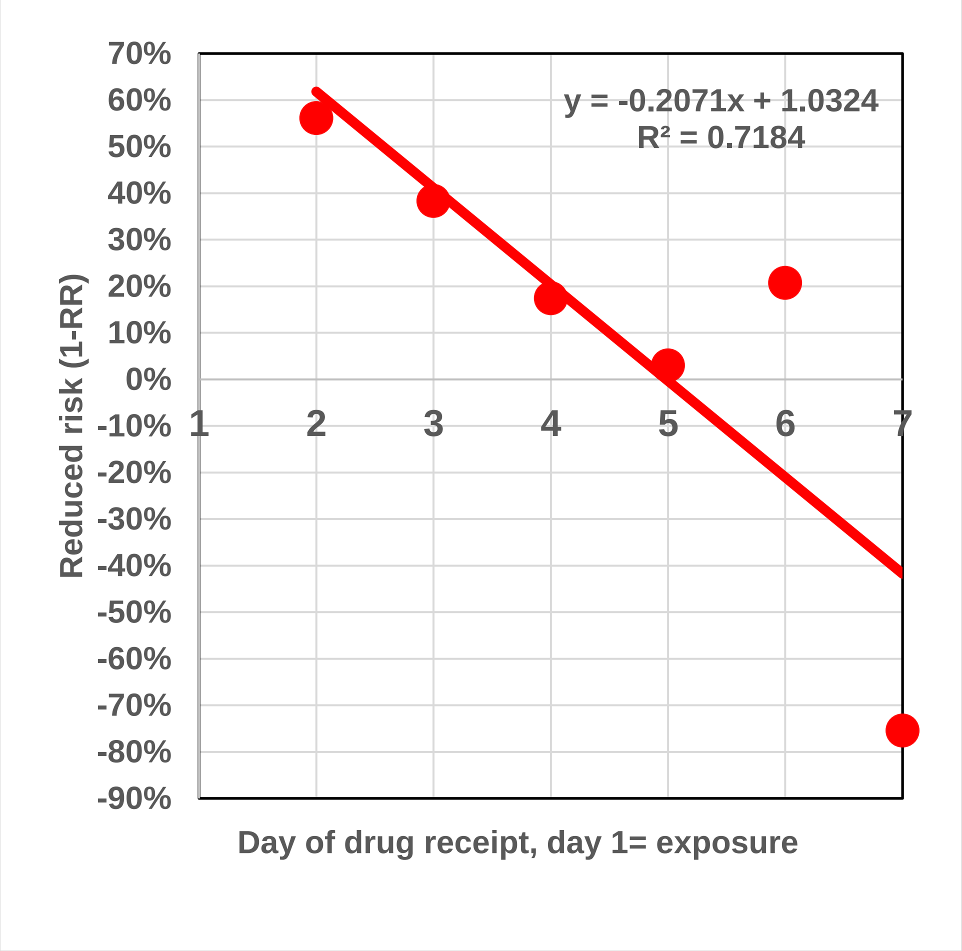

The Days shown are the numbered days on which study drug was received, with Day 1 = day of reported high risk exposure. See note in Table 1 regarding the nomenclature for "days." All comparisons are with the placebo cohort, pooled across all time periods. The Days shown are the numbered days on which study drug was received, with Day 1 = day of reported high risk exposure. See note in Table 1 regarding the nomenclature for "days." All comparisons are with the placebo cohort, pooled across all time periods. A linear regression line was plotted with Microsoft Excel. The p value (2 tailed) for the regression line is 0.033. The 95%CI for rho are: -0.982, -0.115. The 95%CI for the slope of the regression are -38.7 and -2.67 (vassarstats.net).

**Revision History**

The following manuscript revisions have been made:
Version 2: Revisions since version 1 of this manuscript [34] was pre-printed December 2, 2020.

i) Additional comments related to just published full paper version of Barnabas et al., [32] previously cited as abstract version. Discussion of dose and loading dose effects between comparable studies, with added Table 5.
ii) Revision of discussion to reflect age strata revision in PrEP study from pre-print to published versions
We had noted in our pre-printed protocol [25] that the age strata used in the pre-printed companion PrEP study [29] differed from those used in the companion PEP [33] and PET [35] studies. These strata have been revised in the published version [30] to match the other two studies. The pre-printed Version 1 of this manuscript also reflected the earlier age strata, which we now revise and add a comparison (Table S 6) between the PEP and PrEP studies.
iii) Table (Table S 5) added for effect of HCQ by risk level and contact type. Differences in the baseline incidence of Covid-19 and the relationship between contact type, risk level and changes in risk definitions are further discussed in main body and Supplement.
iv) Discussion revised concerning statistical limitations.
v) Comments added regarding remaining need for non-vaccine approaches.
vi) Citation of Mitja [36] study: Some RR values changed to aRR to reflect adjustment noted in Mitja Table 2 and apparent discrepancy with values in Mitja Figure 1.
vii) Table 4: non-stratified totals added
viii) Additional discussion in limitations.
ix) Use of the term “intervention lag” to denote the time between highest risk exposure and receipt of study medication.
x) Updated citations from pre-print to published versions.
xi) Various typos, minor grammatical errors, reduction of word count.
xii) Typo in Luco analysis, p value corrected to 0.0293 from 0.293.
xiii) Verification calculations: minor differences likely due to rounding noted (Supplement)
xiv) Description of selection of strata boundaries for age - expanded and moved from main text. (Supplement)

Versions 3.1-3.4: Revisions since version 2 of this manuscript [34] was pre-printed December 12, 2020.

i) Analysis confirming Watanabe, considering subjects becoming Covid-19 positive after the five-day treatment period (Table S 7). Comment added in discussion.
ii) Abella, Bhattacharya. references and comments removed (to preserve wound count and reference limit)
iii) Yang [37] reference inserted, noting their independent observation regarding the shipping time issue, recently brought to our attention.
iv) Revised discussion of statistical limitations, Mudge et al., reference added.
v) Plain language summary moved to Supplement
vi) CDC reference [38] removed, duplicates information in WHO reference. [39]
vii) Pharmacokinetic study by Boulware group referenced [40] and integrated into discussion of comparable studies.
viii) Pragmatic trial reference replaced with better citation.
ix) Table 5, weight added
x) Typos and word economy, update references and format
xi) Running and extended title
xii) Clarifications and additional analyses to address comments made on medrxiv. These do not affect study results or interpretation.

a. Clarification regarding “day on” vs. “days from” nomenclature added throughout.
b. Clarification of variables used in different versions of the dataset (Supplement), including a newly circulating, but inaccurate revision (10/26).
c. Sensitivity analysis using pooled placebo cohort to compare time-related effects.
d. Sensitivity analysis based on Figure 1 of Nicol et al. [26]

xiii) Title change to “novel, missing, data”

xiv) Highlights added

[38] Centers for Disease Control and Prevention. CDC COVID Data Tracker. 2020. (Accessed Dec 16, at covid.cdc.gov/covid-data-tracker/.)

[39] World Health Organization. WHO Coronavirus Disease (COVID-19) Dashboard. 2021. (Accessed June 11, at https://covid19.who.int/.)

[40] Al-Kofahi M, Jacobson P, Boulware DR, Matas A, Kandaswamy R, Jaber MM, et al. Finding the Dose for Hydroxychloroquine Prophylaxis for COVID-19: The Desperate Search for Effectiveness. Clinical pharmacology and therapeutics. 2020 Apr 29;108:766-9. http://doi.org/10.1002/cpt.1874
